# Longitudinal bidirectional effects between ADHD and education performance: a new RI-CLPM model with instrumental variables

**DOI:** 10.1101/2025.08.06.25333140

**Authors:** Luis FS Castro-de-Araujo, Madhurbain Singh, Daniel Zhou, Philip Vinh, Sydney Kramer, Hermine HM Maes, Michael C Neale

## Abstract

Attention-Deficit/Hyperactivity Disorder (ADHD) is a prevalent neurodevelopmental disorder with significant implications for educational attainment. Previous research has established a bidirectional relationship between ADHD symptoms and academic outcomes, but the underlying mechanisms remain poorly understood. This study employs a novel Random Intercept Cross-Lagged Panel Model (RI-CLPM) incorporating instrumental variables to improve precision of bidirectional effects estimation between ADHD and educational performance. Utilizing a large, longitudinal dataset from the Adolescent Brain and Cognitive Development Study (ABCD Study^®^), we examine the reciprocal influences between ADHD symptoms and academic achievement across multiple time points. Our findings reveal significant within-wave bidirectional effects, with ADHD symptoms negatively impacting subsequent academic attainment, and poor academic outcomes exacerbating ADHD symptoms over time. These results underscore the need for integrated interventions targeting both ADHD symptoms and academic support.

## 1. Introduction

Attention-Deficit/Hyperactivity Disorder (ADHD) is among the most prevalent externalizing disorders in children. Its prevalence estimates range from 7.2% (95%CI 6.7-7.8) to 7.6% (95%CI 6.1–9.4%) in children (Thomas et al., 2015). However, the condition can also affect adolescents and adults (Boomsma et al., 2010; Choi et al., 2022; Larsson et al., 2014; Willcutt, 2012). The cardinal symptoms are inattention, hyperactivity and impulsivity. ADHD is often co-morbid with other psychiatric disorders, like obsessive-compulsive disorder, autism, learning disorders (Fayyad et al., 2017; Sousa et al., 2020), and alcohol use (Demange et al., 2023). According to the DSM-5, symptoms should be present in two or more settings, such as at home, school or work and must impair social, school, or work functioning (APA, 2013). ADHD has been shown to be negatively correlated with academic success (Boomsma et al., 2010; Demontis et al., 2019).

ADHD diagnosis or symptom scores are highly heritable, consistent with a polygenic model of numerous genetic loci, each exerting a small effect (Faraone and Larsson, 2019). Twin studies estimate the broad-sense heritability of ADHD to be about 80% (Chen et al., 2017), while adoption studies show negligible shared environmental impacts (Sprich et al., 2000). A ninefold increase in risk of ADHD was reported in siblings of those affected compared to siblings of controls, with a sibling correlation coefficient of 0.3 (Chen et al., 2008; Faraone and Larsson, 2019). Heritability appears equal across sexes (Faraone and Larsson, 2019; Larsson et al., 2014), and the genetic correlation for ADHD across sex is nearly one (Taylor et al., 2016) although the prevalence ratio of males to females varies from 2:1 to 2.5:1 depending on instrument used in the assessment (Hinshaw et al., 2022; Sousa et al., 2020).

Educational attainment (EA) is a complex trait influenced by genetic, environmental, and socio-cultural factors. Epidemiologically, EA is often measured by the highest level of education completed and is a strong predictor of various life outcomes, including income, health, and longevity (Murray, 2013). Genetic epidemiology studies have increasingly highlighted the heritable nature of EA, with twin and family studies estimating heritability to be around 40% with a relatively large non-shared environment variance of 36% (Branigan et al., 2013). Genome-wide association studies (GWAS) have identified numerous genetic loci associated with EA, suggesting that it is a polygenic trait influenced by many small-effect genetic variants (Lee et al., 2018). Moreover, gene-environment interactions play a significant role in shaping educational outcomes, with socio-economic status and educational policies acting as critical moderators (Rimfeld et al., 2018).

Recent evidence supports a bidirectional causal relationship between ADHD symptoms and educational attainment (EA) using three distinct methodologies: Demange et al. 2024 reports a large two-sample Mendelian Randomization (MR) study between EA and many psychiatric phenotypes, including ADHD. In the same paper they report results from within-sibship analysis (Demange et al., 2024). Using the Dutch national registry data they observed a mean prevalence of around 1% across all years of the study, but a peak at around 3% when participants reach ten years of formal education, decreasing thereafter. Prevalence was slightly higher in males. They found indication of bidirectional causation between EA and ADHD using MR, with IVW OR of 0.73 (P < 0.004) for the influence of EA on ADHD and IVWβ=-0.38 (P < 0.004) for the reverse effect, suggesting possible bidirectional causal relationship.

Dardani et al. (2021) reported results from a two-sample MR where they found a significant negative effect of ADHD on EA (IVWβ= −.05 [95%CI −.08, −.02]) and of EA on ADHD symptoms (OR .33 [95%CI .26, .43]), independent of cognitive ability. Likewise, Michaëlsson et al., (2022) reports a IVW OR of .33 [95%CI .21, .50] of EA on ADHD and a more modest IVWβ −0.09 [95%CI −.14, −.05] from ADHD on EA. However, sensitivity analyses with distinct MR methods were not consistent with the latter result.

Previous results are limited to report results in adults, given all GWAS are in that population. In the youth, however, the available metric is their performances at school. Using family-based causal inferential methods, Castro-de-Araujo et al. (2024) reported significant bidirectional causal relationships between an ADHD symptom scale and education performance (EP, grades from last school year) in adolescents from the Adolescent Brain and Cognitive Development Study (ABCD Study^®^). They reported −0.27 [95%CI −0.412, −0.124], −0.26 [95%CI: −0.333, −0.184] and −0.07 [95%CI: −0.261, 0.122] ADHD to EP coefficients using Direction of Causation models in European (EUR), American (AMR), and African (AFR) ancestries, respectively. The reverse relationship reported effect sizes were respectively −0.73 [95%CI −1.11, −0.35], −0.9 [95%CI: −1.11, −0.839], and −0.06 [95%CI: −0.449, 0.334], the last was not significant, likely due to the small sample size of the AFR group. Results from instrumental variable analysis in twins, using the Polygenic Scores (PS) for each phenotype as an instrumental variable in the EUR ancestry group, resulted in significant effects: −0.25 [95%CI −0.34,-0.15] from ADHD symptoms to school grades and −0.693 [95%CI −0.9512,-0.4351] in the reverse direction. Finally, results from a bidirectional instrumental variables method in twins resulted in −0.86 [95%CI −0.97,-0.73] forward effect and −0.5 [95%CI −0.58,-0.41] in the reverse direction (Castro-de-Araujo et al., 2024). The triangulation of these methodologies suggests an underlying causal relationship between ADHD and EP in both directions. All these studies are cross-sectional. These findings indicate that ADHD not only hinders educational outcomes, but also that addressing learning challenges — potentially through increased support from parents, schools, or teachers — may positively influence the disorder’s clinical trajectory.

Understanding the causal link of ADHD and education performance over childhood and adolescent development is of major importance to identifying optimal timing for intervention. Cross-Lagged Panel Models (CLPM) have been helpful to elucidate temporal dependencies between phenotypes (Kenny, 1975; Selig and Little, 2012). However, classic CLPM is informative about causal links in the sample, not at the individual level. Hamaker et al. (2015) extended these models with random intercepts (RI) and unlike conventional CLPM, which assumes that the observed variables at each time point are solely influenced by their previous states and the previous states of other variables, the RI-CLPM incorporates random intercepts to account for stable, trait-like differences between individuals. This model effectively separates the within-person (state-like) fluctuations from the between-person (trait-like) differences, providing a more nuanced understanding of the temporal dynamics. Similarly, Singh et al. (2023) extended CLPM to incorporate instrumental variables (IV-CLPM) improving estimation when measures are too distant apart. By incorporating IVs, which are variables that influence the endogenous predictors, but are not directly related to the outcome variables except through these predictors, the model provides a more robust framework for disentangling the bidirectional influences between variables.

The inspection of causal relationships as children age may reveal the optimum moment in development before which intervention will be more effective. Importantly, methods that can assess these causal links bidirectionally are preferred, because they provide information on whether interventions should aim at the child’s ADHD symptom severity or their educational environment, or both. Given the current evidence on these phenotypes, it is reasonable to think that improvements in either or both ADHD and the educational environment should provide improvement of clinical course by reducing ADHD symptoms and enhancing EA. The aims of this study are to develop a new model that incorporates both instrumental variables and random intercepts with CLPM (IV-RI-CLPM); (ii) evaluate the effect of increasing lag times on its power and estimates, comparing it to CLPM, RI-CLPM and IV-CLPM; (iii) provide a substantive application seeking whether the causal links between ADHD and education performance found previously are stable over three follow-up waves in the European ancestry subsample of the Adolescent Brain Cognitive Development (ABCD®) study. In what follows, the substantive application will be presented first and simulations second.

## 2. Methods

### 2.1. Data source

Data were sourced from the ABCD study, release v5.1 (Haist and Jernigan, 2023), a comprehensive, long-term study of brain development and child health of over 10,000 children across the U.S. The research encompasses multiple domains, such as neuroimaging, genetics, cognitive performance, mental health, substance use, and environmental influences (Maes et al., 2022). The phenotypes of interest were both present in EUR ancestry participants at follow-up year 2 (n=5515), 3 (n=5198), and 4 (n=2519). Data for follow-up year 4 were incomplete at the time of analysis, comprising about half of the study participants.

### 2.2 Instruments and polygenic score calculation

The Child Behavior Checklist (CBCL) ADHD subscale was used to assess ADHD symptoms and their longitudinal changes. This caregiver-reported subscale encompasses items on inattention, hyperactivity, and impulsivity (Achenbach, 2001). The CBCL ADHD subscale is extensively used to evaluate behavioral issues in children and adolescents and aligns well with DSM criteria for ADHD (Achenbach, 2013). When compared to clinical evaluations, the CBCL has demonstrated a sensitivity of 0.77 and a specificity of 0.73 for ADHD diagnosis (Chang et al., 2016). DSM-oriented CBCL scales generally show a high Cronbach’s alpha of 0.92, while the CBCL ADHD subscale has an area under the curve of 0.75 for diagnostic accuracy (Jiang et al., 2023). The 14-level sum-score variable was transformed into a 6-level ordinal variable for model fitting.

The second phenotype examined was the youths’ school grades from the previous year, as reported by parents. This variable was coded so that 1 represented an A+ and 12 an F. It was then reversed so that higher values indicated better performance and was included in the models as a 7-category ordinal variable.

PSs were derived from the latest GWAS for each phenotype. ADHD liability PS were estimated using summary statistics from Demontis et al. (2023). PS for educational attainment were calculated using summary statistics from Okbay et al. (2022). Due to the lack of GWAS summary statistics for non-European ancestry groups, the PSs were computed in ABCD samples of European ancestry only, ancestry assignment was provided by the study (Haist and Jernigan, 2023). For this reason, only participants of European ancestry were used for these analyses. Quality Control (QC) on the GWAS summary statistics followed standard guidelines (Choi et al., 2020), filtering out SNPs with minor allele frequencies (MAF) <0.01, INFO scores <0.8, duplicate SNPs, and ambiguous variants (e.g., A-T or G-C). QC of genotypic data in the target ABCD sample also adhered to standard protocols, excluding SNPs with a MAF <0.01, Hardy-Weinberg-Equilibrium Fisher’s Exact Test p-values <10^-6^, and missingness rates >0.01. Individual samples with genotype missingness rates >0.01 were also excluded. PRS-cs v 1.1.0 (Ge et al., 2019) was employed to infer posterior SNP effect sizes, which were then used to compute individual-level PS in the target ABCD study samples using PLINK 1.9 (Purcell et al., 2007). The phenotype was regressed on the PRS for the full data set used in the analyses, and it was found an adjusted R^2^ of 0.02 with an F-statistic of 37.3 for CBCL ADHD, and an adjusted R2 0.02 and F-statistic 35.4 for grades.

### 2.3 Covariates

Sex was incorporated as a covariate. In order to control for population stratification, the first six principal components (PCs) were included as covariates. These were within-ancestry PCs calculated in-house for the European sample from the ABCD study® using PC-AiR (Conomos et al., 2015). Only six were included because the last PCs were not correlated to either phenotype.

### 2.4. Trans-diagnostic validation

After the IV-RI-CLPM was fit to the data, the factor scores of the ADHD random intercept were calculated (variance parameter rix, see below) by passing the model to umxFactorScores, which in turn generate scores for all latent variables within the model. These scores were then merged with the original data set and polychoric correlations with Urgency-Premeditation-Perseverance-Sensation Seeking (UPPS) scale domain scores were estimated.

### 2.5. Data analysis

R version 4.1.1 (https://cran.r-project.org/) was used for all subsequent analyses, utilizing the OpenMx v2.20.7 and umx v4.19.0 packages for structural equation modeling (SEM) (Bates et al., 2019; Neale et al., 2016). Full information maximum likelihood (FIML) with the SLSQP optimizer was used for parameter estimation. Model identification was confirmed using the OpenMx utility mxCheckIdentification (Hunter et al., 2021). Due to the high computational demands of treating both phenotypes as ordinal categorical variables, analyses were performed in 23 CPUs from the VIPBG cluster (https://vipbg.vcu.edu/resources/computational-facilities). A comprehensive list of loaded R packages for each analysis is available in the repository for this study (https://github.com/lf-araujo/2024-RI-CLPM). Descriptive statistics are presented in Table 1. An alpha level of 0.05 was used for all tests, with a greater emphasis placed on effect size over significance, given the extensive sample size of ABCD (Dick et al., 2021; Owens et al., 2021).

**Table 1.** Main demographic characteristics of the European ancestry ABCD sample, stratified by study wave. The CBCL ADHD scores, UPPS scale scores, and the students’ grades in the previous year were treated as ordinal variables.

|  | Follow-up<br>year 2 | Follow-up<br>year 3 | Follow-up<br>year 4 |
| --- | --- | --- | --- |
| n | 5515 | 5198 | 2519 |
| CBCL ADHD score (%) |  |  |  |
| 0 | 1653 (30.0) | 1523 (29.3) | 1008 (40.0) |
| 1 | 718 (13.0) | 679 (13.1) | 431 (17.1) |
| 2 | 419 ( 7.6) | 455 ( 8.8) | 264 (10.5) |
| 3 | 384 ( 7.0) | 387 ( 7.4) | 236 ( 9.4) |
| 4 | 305 ( 5.5) | 315 ( 6.1) | 170 ( 6.7) |
| 5 | 233 ( 4.2) | 226 ( 4.3) | 107 ( 4.2) |
| 6 | 164 ( 3.0) | 169 ( 3.3) | 91 ( 3.6) |
| 7 | 130 ( 2.4) | 111 ( 2.1) | 76 ( 3.0) |
| 8 | 82 ( 1.5) | 84 ( 1.6) | 38 ( 1.5) |
| 9 | 62 ( 1.1) | 59 ( 1.1) | 35 ( 1.4) |
| 10 | 41 ( 0.7) | 38 ( 0.7) | 22 ( 0.9) |
| 11 | 32 ( 0.6) | 24 ( 0.5) | 16 ( 0.6) |
| 12 | 28 ( 0.5) | 20 ( 0.4) | 12 ( 0.5) |
| 13 | 11 ( 0.2) | 18 ( 0.3) | 8 ( 0.3) |
| 14 | 8 ( 0.1) | 11 ( 0.2) | 3 ( 0.1) |
| NA | 1245 (22.6) | 1079 (20.8) | 2 ( 0.1) |
| Grades last year (%) |  |  |  |
| 1 | 8 ( 0.1) | 16 ( 0.3) | 18 ( 0.7) |
| 2 | 18 ( 0.3) | 30 ( 0.6) | 33 ( 1.3) |
| 3 | 35 ( 0.6) | 27 ( 0.5) | 30 ( 1.2) |
| 4 | 68 ( 1.2) | 94 ( 1.8) | 59 ( 2.3) |
| 5 | 121 ( 2.2) | 128 ( 2.5) | 74 ( 2.9) |
| 6 | 179 ( 3.2) | 185 ( 3.6) | 94 ( 3.7) |
| 7 | 268 ( 4.9) | 275 ( 5.3) | 145 ( 5.8) |
| 8 | 581 ( 10.5) | 553 ( 10.6) | 262 ( 10.4) |
| 9 | 737 ( 13.4) | 636 ( 12.2) | 308 ( 12.2) |
| 10 | 954 ( 17.3) | 889 ( 17.1) | 391 ( 15.5) |
| 11 | 1608 ( 29.2) | 1539 ( 29.6) | 712 ( 28.3) |
| 12 | 802 ( 14.5) | 698 ( 13.4) | 361 ( 14.3) |
| NA | 136 ( 2.5) | 128 ( 2.5) | 32 ( 1.3) |
| Sex = M (%) | 2912 (52.8) | 2750 (52.9) | 1339 (53.2) |
| Age (mean (SD)) | 12.04 (0.67) | 12.93 (0.65) | 14.13 (0.69) |
| UPPS negative urgency<br>(mean (SD)) | 7.66 (2.26) |  | 8.18 (2.34) |
| UPPS lack of planning (mean<br>(SD)) | 7.84 (2.19) |  | 8.03 (2.13) |
| UPPS sensation seeking<br>(mean (SD)) | 9.68 (2.68) |  | 10.21 (2.67) |
| UPPS positive urgency<br>(mean (SD)) | 7.15 (2.55) |  | 7.64 (2.56) |
| UPPS lack of perseverance<br>(mean (SD)) | 6.85 (2.22) |  | 7.36 (2.35) |

### 2.6. Model specification

The McArdle and McDonald Reticular Action Model (RAM) was used to specify the models via path syntax in umx (Bates et al., 2019; Neale et al., 2016). The new model presented here (IV-RI-CLPM, Figure 1) adds elements to the IV-CLPM model (Singh et al., 2023). Notably, the instrument effect on X Y: PSx and PSy, respectively (Figure 1) and the within-wave causal paths (e and f parameters, Figure 1). In the IV-RI-CLPM, the residual variances of p and q are innovations (u and v, Figure 1). In the final model residual variances are equated across waves for p (u) and for q (v), as well as their correlations (uv) (yellow, orange and purple double-headed arrows in Figure 2). The residual correlations between X and Y allows and their innovations (u, v, and uv) represent background confounding in cross-sectional IV regression. The presence of PSx and PSy identifies the within-wave causal paths, and the background confounding correlation in classic MR (Figure 1). Covariates were included in the SEM model as definition variables, therefore the distributional properties of the covariates do not affect the estimation.

**Figure 1:**
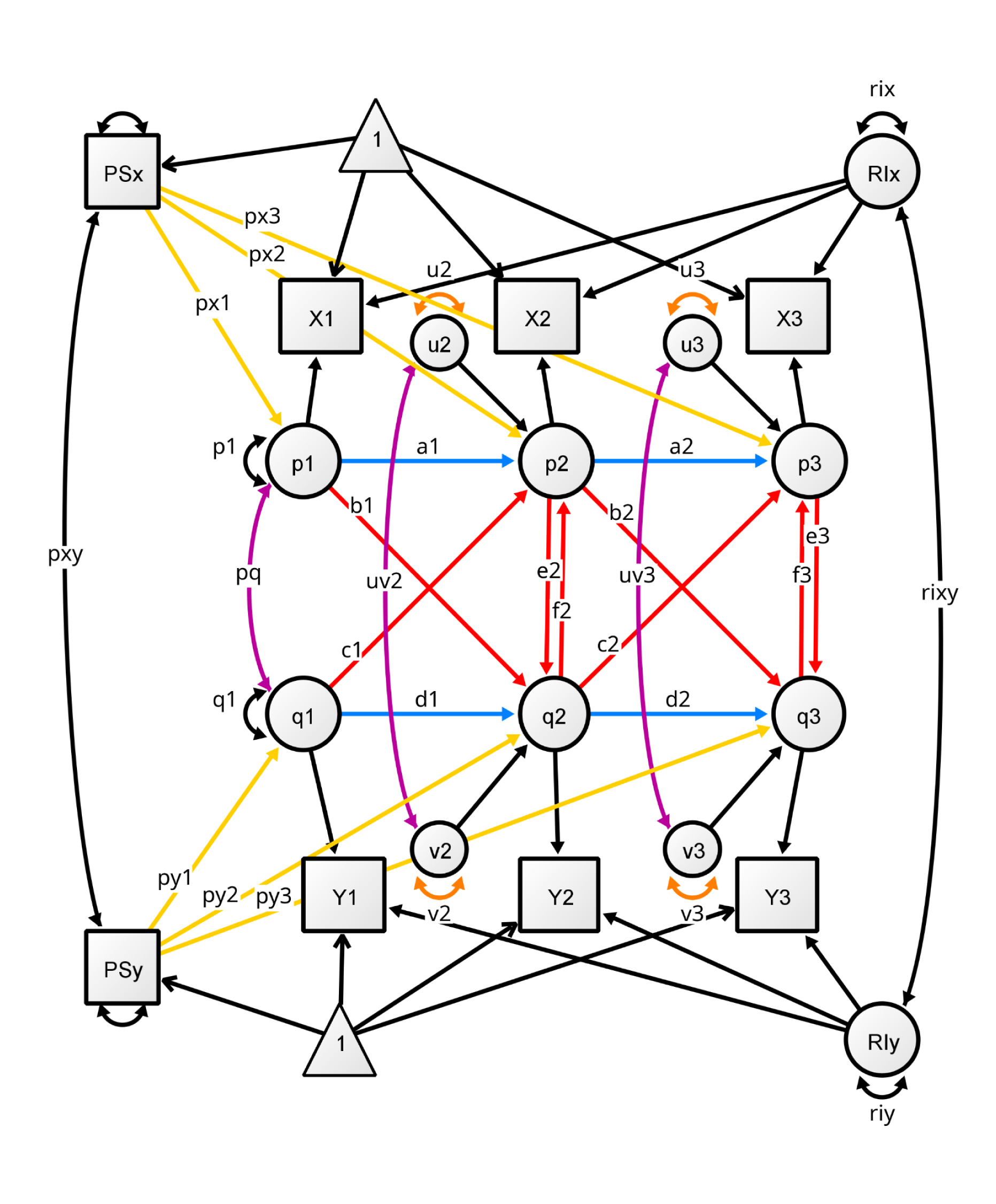
Model specification. Note: px, path from instrument to observed; p, X variances; q, Y variances; u, innovation variances for X; v, innovation variances for Y; uv, correlation between us and vs; RIx, random intercept for X; RIy, random intercept for Y; rixy correlation between random intercepts. No equality constraints were used in the simulations.

**Figure 2:**
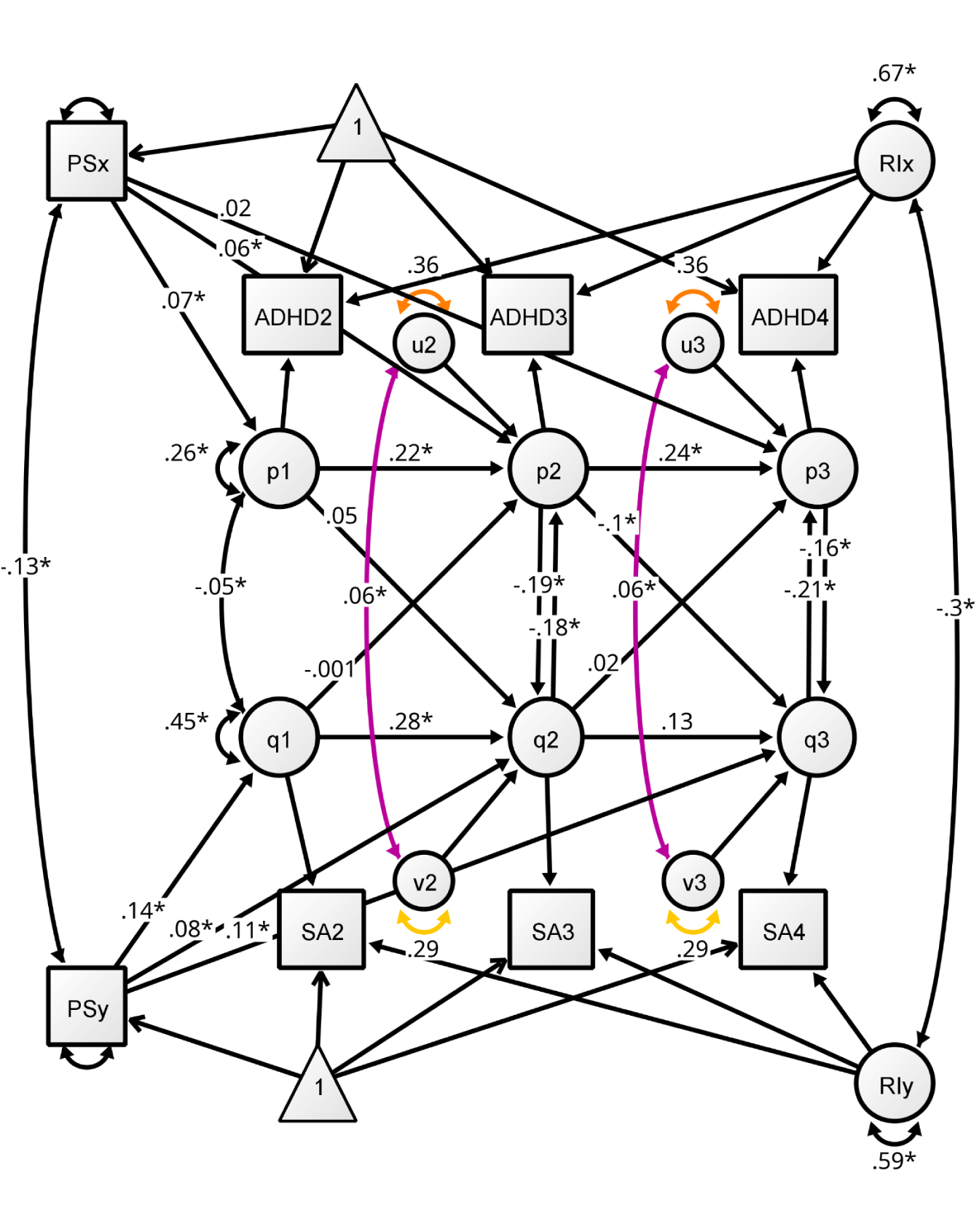
Model estimation. Instrumental variables random intercepts CLPM. The model is identified as depicted, variances for innovations are equal across waves (orange and yellow double-headed arrows, u & v), innovation correlations are also equal across waves (purple). The model comprises 5 main elements: (A) the observed variables shown as rectangles; (B) the between-person variances (variances for RIy and RIx); (C) the instrumental variables (polygenic scores, PSx and PSy); (D) the means for all observed (not shown); and (E) the random intercepts (RIy and RIx). Fixed paths are marked with 1. Values are point estimates in the ADHD and education performance analysis. In this analysis the observed variables were treated as ordinal in the latent liability scale, in this case OpenMx does not generate fit indices. The fit for the all continuous analysis was: χ²(10) = 41.92, p < 0.001; CFI = 0.997; TLI = 0.991; RMSEA = 0.024. The estimation was controlled for sex and first 6 principal components of ancestry.

### 2.7. Simulation procedures

A limitation of conventional CLPM is that as lag increases, power to detect the causal effects reduces. Simulations were performed to check what the addition of IVs and RIs to conventional CLPM results in terms of power and precision. We simulated time-series data with autoregression and reciprocal causal effects between two variables, X and Y (Figure 1), to compare RI-IV-CLPM with RI-CLPM (Hamaker et al., 2015). The simulated data included two instrumental variables (IVs): PSx directly associated with X, and PSy directly associated with Y, at each time point. The data-generating model is depicted in Figure 1.

Data were generated for 20 time points using exact simulation of the expected covariance matrix given the parameter values listed in Table 2. We declared the model to be stationary as beginning at time t occasion when the cross-sectional correlations between X and Y at time t differed by less than 0.0002 at from those at timepoint t+1. This closely approximates stationarity which is declared when means, variances, and covariances of the variables do not change over time. Data were simulated for N = 1000 employing the mvrnorm() function in the MASS package (Venables et al., 2002), with the option empirical = TRUE. This approach ensures that the covariance matrix of the simulated data exactly matches the predicted covariance matrix (van der Sluis et al., 2008). Under these conditions, twice the difference in negative log-likelihood of a model where a parameter is free versus fixed to zero can be used as a non-centrality parameter (NCP) from which statistical power can be calculated. Plots for the within-occasion correlations between X and Y are shown in Figure 4 and Figure 9. IV-RI-CLPM and RI-CLPM were considered stationary beyond time-point 12. Notably, when the auto-regressive paths were smaller (a and d) the model reached stationarity at an earlier time-point. Not stationary data points, i.e., before T = 12, were discarded.

**Figure 4.**
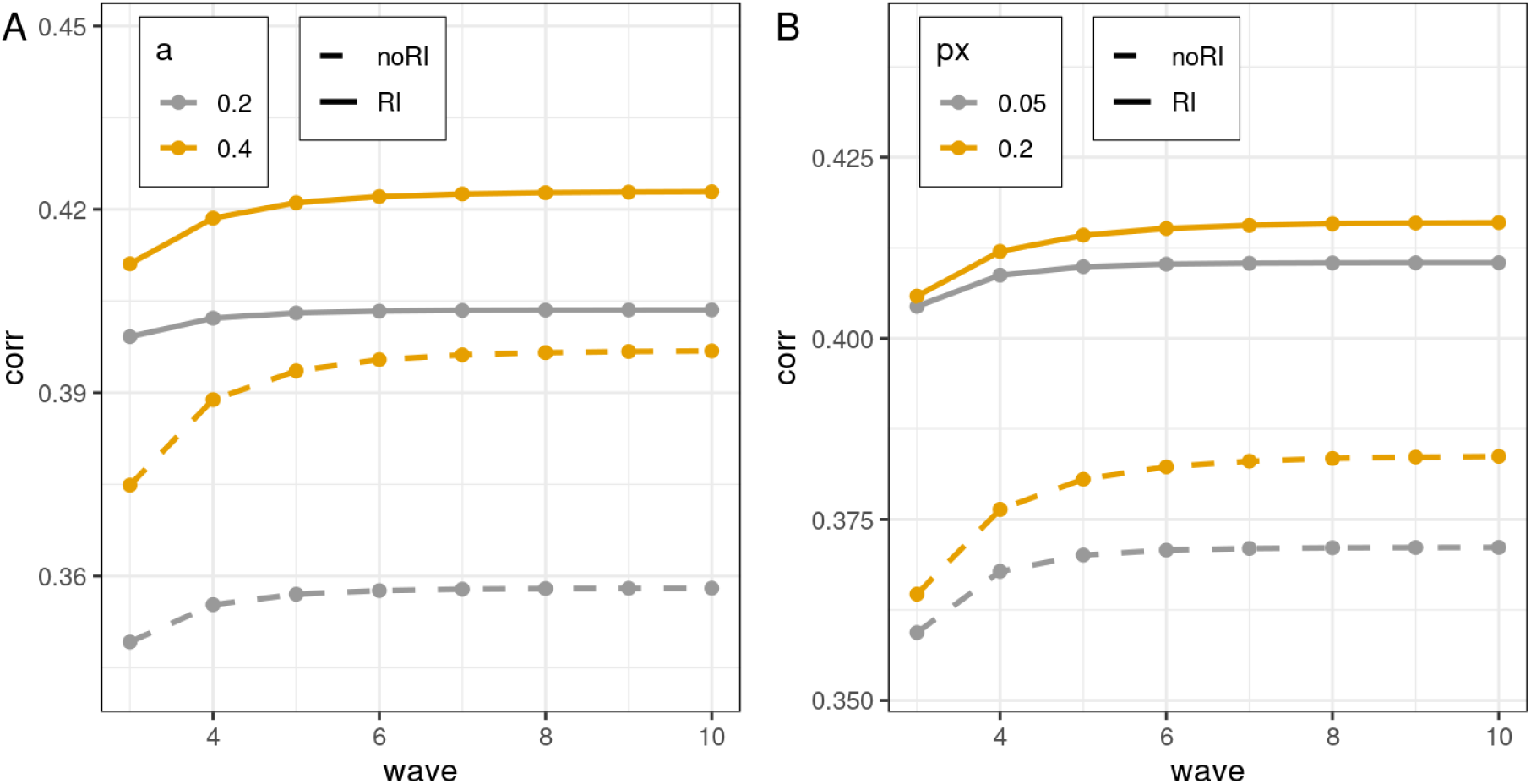
Stationarity - IV-RI-CLPM. Correlation between Xt and Yt, where t is the study wave up to 10. With the extension in time of the model, the correlation tends to stabilize. The correlation is plotted by parameter values (a, px) and linetype representing the presence or absence of random intercepts (RI, noRI). NoRI is equivalent to Singh et al. (2023) IV-CLPM. px, the effect size of the path from the instrument to X; a, the effect size of the auto-regressive path. Increasing the auto-regressive path or the effect size of the instrument delays the stationarity of the model.

**Figure 9.**
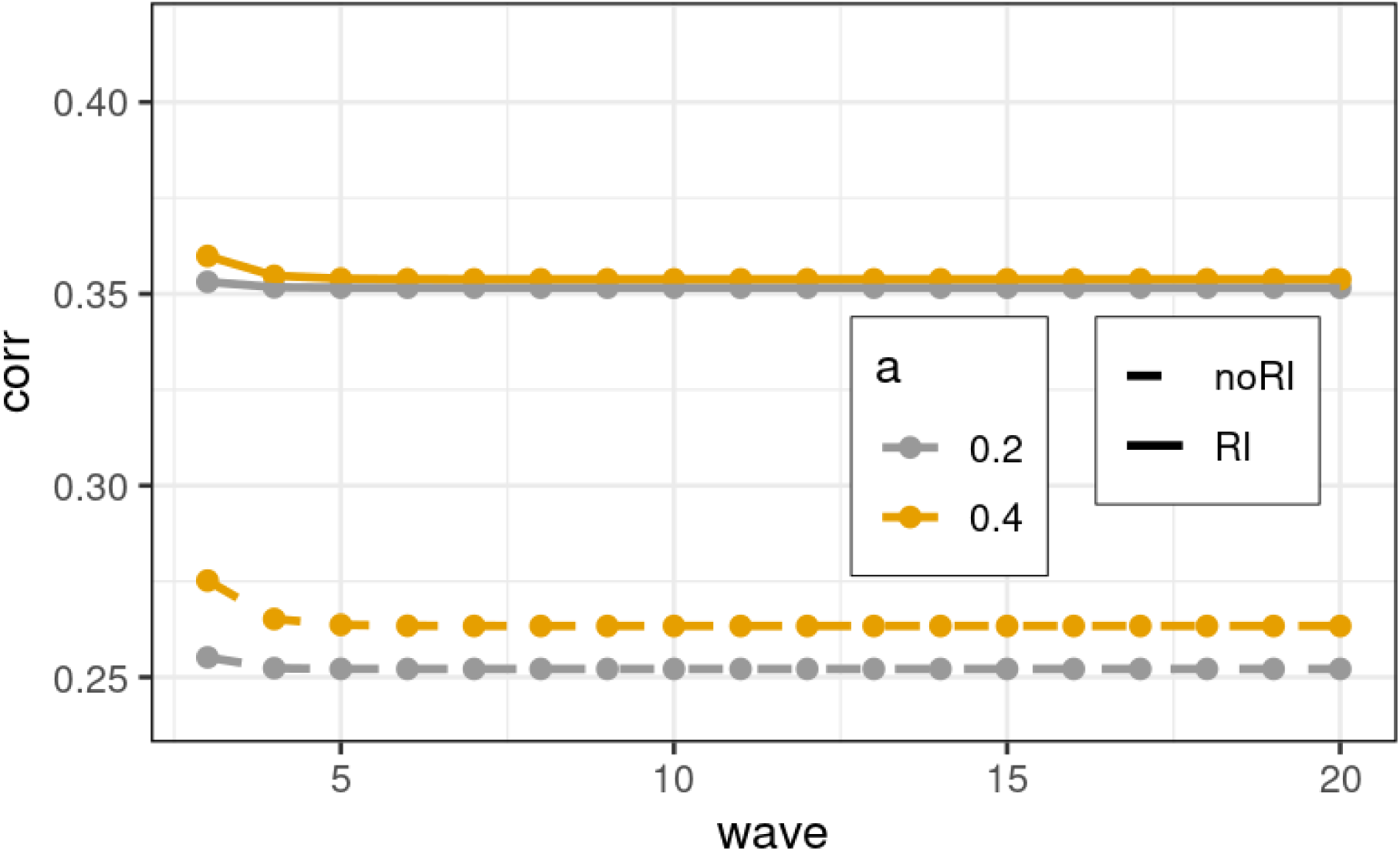
RI-CLPM stationarity. Correlation between Xt and Yt, where t is the study wave up to 20. With the extension of the model, the correlation stabilizes. This model reaches stationarity at wave 10 in all cases listed. The correlation is plotted by parameter values (a, px) and faceted by the presence or absence of random intercepts (RI, noRI). NoRI is equivalent to Singh et al. (2023) IV-CLPM. px, instrument to X path coefficient; arp, auto-regressive path coefficient. Increasing the auto-regressive path or the effect size of the instrument delays the stationarity of the model similar to what was found in the model proposed (IV-RI-CLPM, Figure 4).

**Table 2.**
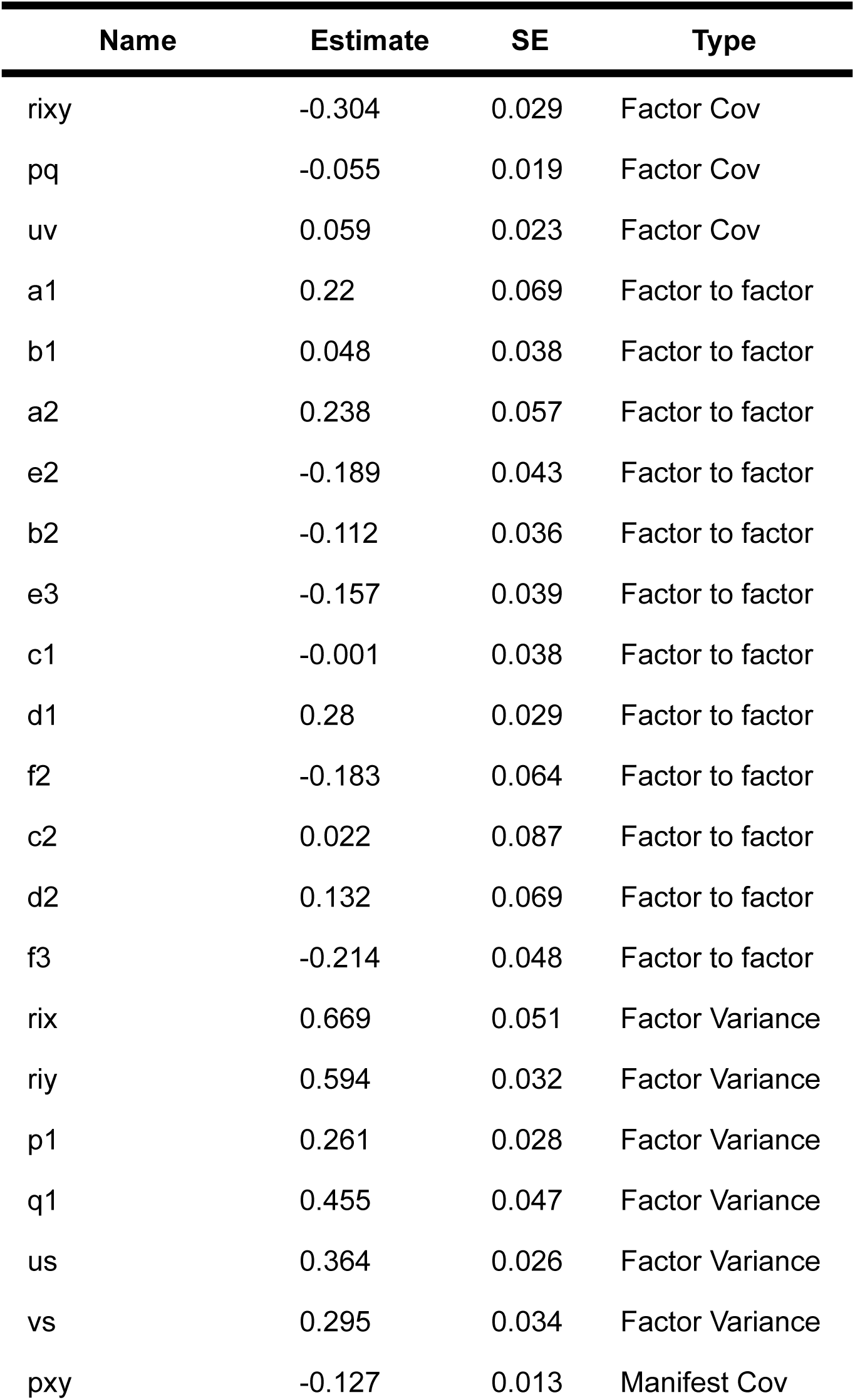
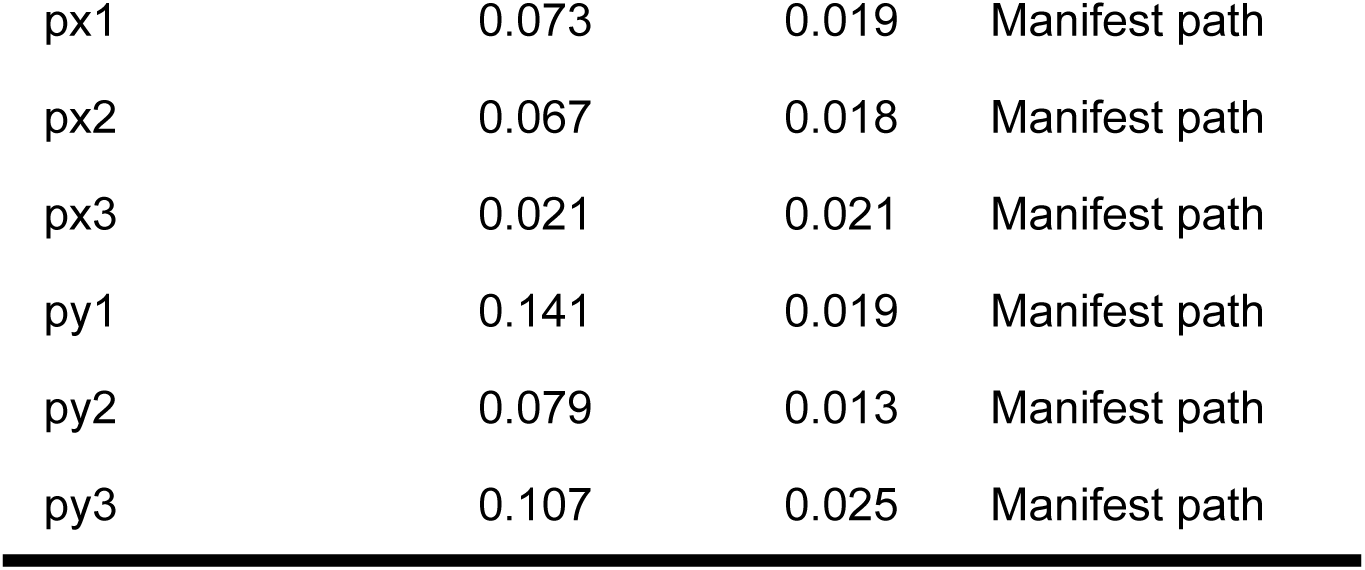
Parameter estimates in the IV-RI-CLPM applied to the ABCD study data. PSx, path from instrument to observed; p, paths from X variances; q, paths from Y variances; us, innovations variances for X equated across waves; vs, innovations variances for Y equated across waves; uv, correlation between us and vs also equated across waves; xir, random intercept for X; yir, random intercept for Y; xir_with_yir correlation between random intercepts. See in Figure 2.

Multiple datasets were simulated using a factorial design where parameter values are levels and each parameter are factors (Table 3). This allows for the examination of the impact of population parameters on the changes in causal estimates across time intervals. The parameter levels were chosen to be consistent with modest-sized regressions of behavioral and psychiatric traits on exogenous IVs (e.g., polygenic scores) (Castro-de-Araujo et al., 2023).

**Table 3:** Simulation factorial design for the exact data simulations. Each parameter is a factor, some parameters have two true values, or levels in the factorial design. Simulation sample size for IV-RI-CLPM is 2,432, 100% convergence; and for CLPM was 608, 100% convergence.

| Parameter name | Type | Values |
| --- | --- | --- |
| px | Instrument effect on exposure | 0.05, 0.2 |
| b | Cross-lagged path (X to Y) | 0, 0.05 |
| c | Cross-lagged path (Y to X) | 0.1 |
| e | Within-wave causal path (immediate, X to Y) | 0.1, 0.2 |
| f | Within-wave causal path (immediate, Y to X) | 0.1 |
| a | Auto-regressive $X_t$ to $X_{t+1}$ | 0.2, 0.4 |
| d | Auto-regressive $Y_t$ to $Y_{t+1}$ | 0.4 |
| rixy | RI's correlation | 0, 0.5 |
| rix, riy | Between-person variance, RI variance (for both X and Y) | 0.5, 0.6 |
| u, v | Within-person variance, X and Y variances | 0.2, 0.3 |
| p1,q1 | Correlation between X and Y residuals at wave one | Same value as u,v |

After the introduction of a lag, the NCP (or difference in -2lnL) was obtained from the likelihood ratio test of freely estimating the distal path between the last two occasions versus fixing it to zero. Notice that both IV-RI-CLPM and RI-CLPM require at least three time-points for model identification. The results were plotted in Figure 8 for IV-RI-CLPM and in Figure 12 for RI-CLPM. Since with the introduction of a lag, the within-wave causal paths exist immediately before and after the lag, for disambiguation they will be called proximal path 1 and proximal path 2, respectively. The cross-lagged path will be called the distal path to remind the reader that in the simulation the lag is increasing.

**Figure 8.**
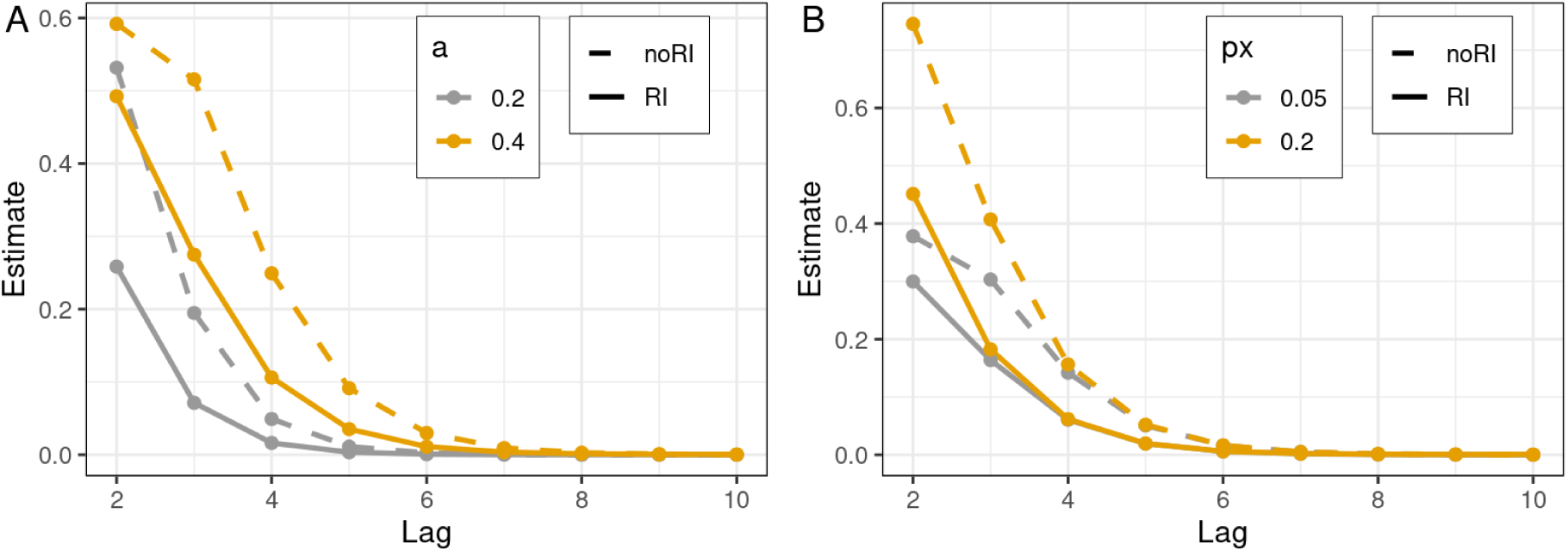
IV-RI-CLPM Power. Power to reject that the distal path from X to Y equals zero as the lag increases (assuming stationarity). The x-axis represents the length of the lag. The y-axis represents the NCP of the LRT with hypothesis of rejecting distal path not equal zero. px, instrument to X path coefficient; arp, auto-regressive path coefficient; Notice the larger arp is, the higher is the power and that the no RI version has higher power than the version with RI.

**Figure 12.**
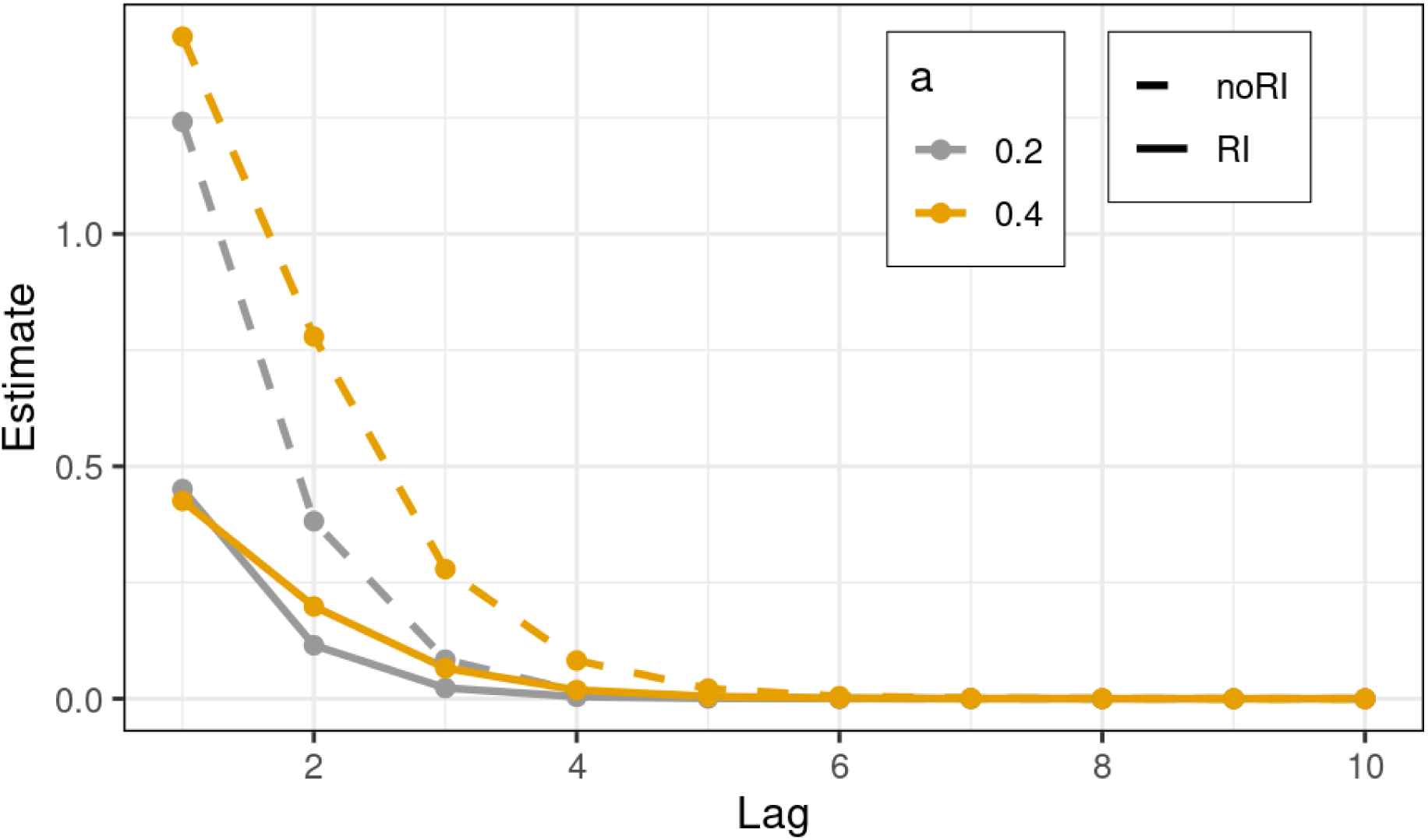
RI-CLPM power. Power to reject that the last distal path from Xt to Yt+lag equals zero as the lag increases (assuming stationarity). The x-axis represents the length of the lag. The y-axis represents the NCP of the LRT with hypothesis of rejecting distal path not equal zero. arp, the effect size of the auto-regressive path. RI-CLPM has less power than the more the conventional CLPM specification (noRI).

## 3. Results

### 3.1. Model fit

The demographic characteristics of the ABCD sample, as presented in Table 1, reveal a decrease in sample size across the study waves (follow-up year 2, follow-up year 3, and follow-up year 4). The sample size decreases from 5515 in follow-up year 2 (flw2) to 2519 in the follow-up year 4 (flw4) due to the fifth release of the ABCD study being incomplete at that occasion, comprising about half of the participants. Additionally, the reduction is in part due to attrition and the Covid-19 pandemic, which started at the end of flw2 and progressed through flw4. The CBCL ADHD scores indicate that the percentage of participants with a score of 0 increases from 30.0% in flw2 to 40.0% in flw4. Conversely, the percentage of participants with higher scores (e.g., score of 3 or above) also shows a slight increase. The majority of students last year grades were scored between 9 and 11, indicating relatively stable academic performance across waves. The sample is predominantly male, with a slight increase in the percentage of males from 52.8% in flw2 to 53.2% in flw4. The mean age of participants is 12.04 years in flw2 and 14.13 years in flw4. The UPPS scale scores showed a slight increase between flw2 and flw4. There was no data for flw3 available.

The observed variables were included as ordinal variables, and the effect reported on the latent liability scale. The covariates were included as definition variables and, unfortunately, OpenMx does not report fit indices in this case. The model was fit with variables treated as continuous, and the fit indices obtained (χ²(10) = 41.92, p < 0.001; CFI = 0.997; TLI = 0.991; RMSEA = 0.024) show good fit (Figure 2). Estimates in Figure 2 are not standardized.

Table 2 presents the parameter estimates from the IV-RI-CLPM of CBCL ADHD and EP. Notable findings include significant negative covariances between the traits at the first time point, between the PSs for both traits, between the RIs for both traits, and positive significant auto-regressive paths for ADHD (0.22 to 0.24) and for EP (0.28 to 0.13). This suggests grades became less stable over time, possibly due to attrition; and that the phenotypes are correlated across time both at state-like and trait-like levels. The only significant cross-lagged path was the causal path from ADHD at Follow-up Year 3 to EP at Follow-up Year 4. All within-wave causal paths were negative and significant. These findings suggest a bidirectional causal effect between EP and ADHD scores.

Only one cross-lagged path was significant, from ADHD to EP −0.112 [95%CI −0.183,-0.042]. As CLPM fails to detect causal paths as lag increases (this will be seen in simulation), maybe the one-year lag for measuring either ADHD CBCL scores or grades is too long for these models to detect the causal effect of state-like fluctuations. These results highlight the complex interplay between ADHD symptoms and EP over time, controlled for sex and the first 6 PCs.

### 3.1. Validation with impulsivity

Impulsivity is genetically associated with ADHD (Lahey et al., 2022) and drug experimentation (Sanchez-Roige et al., 2023). We expect that factor scores derived from the ADHD scores random intercept should be predictive of impulsivity, and showing correlation between these scores and measures of impulsivity in ABCD provides validation that the ADHD trait-like variability with a known associated condition. With this hypothesis, we obtained FIML polychoric correlations between UPPS domain scores and the factor scores calculated from the ADHD symptoms random intercept (rix in Figure 3). It was found that the derived scores have shown varying levels of correlation depending on impulsivity dimensions measured by UPPS in flw2 (Figure 3). I Higher correlation was found between the RI scores scores and lack of perseverance 0.26 (SE 0.01), followed by lack of premeditation (0.19 [SE 0.01]), negative urgency (0.15 [SE: 0.01]), positive urgency (0.14 [SE: 0.01]), and sensation seeking (0.05 [SE 0.01]) (Figure 3).

**Figure 3:**
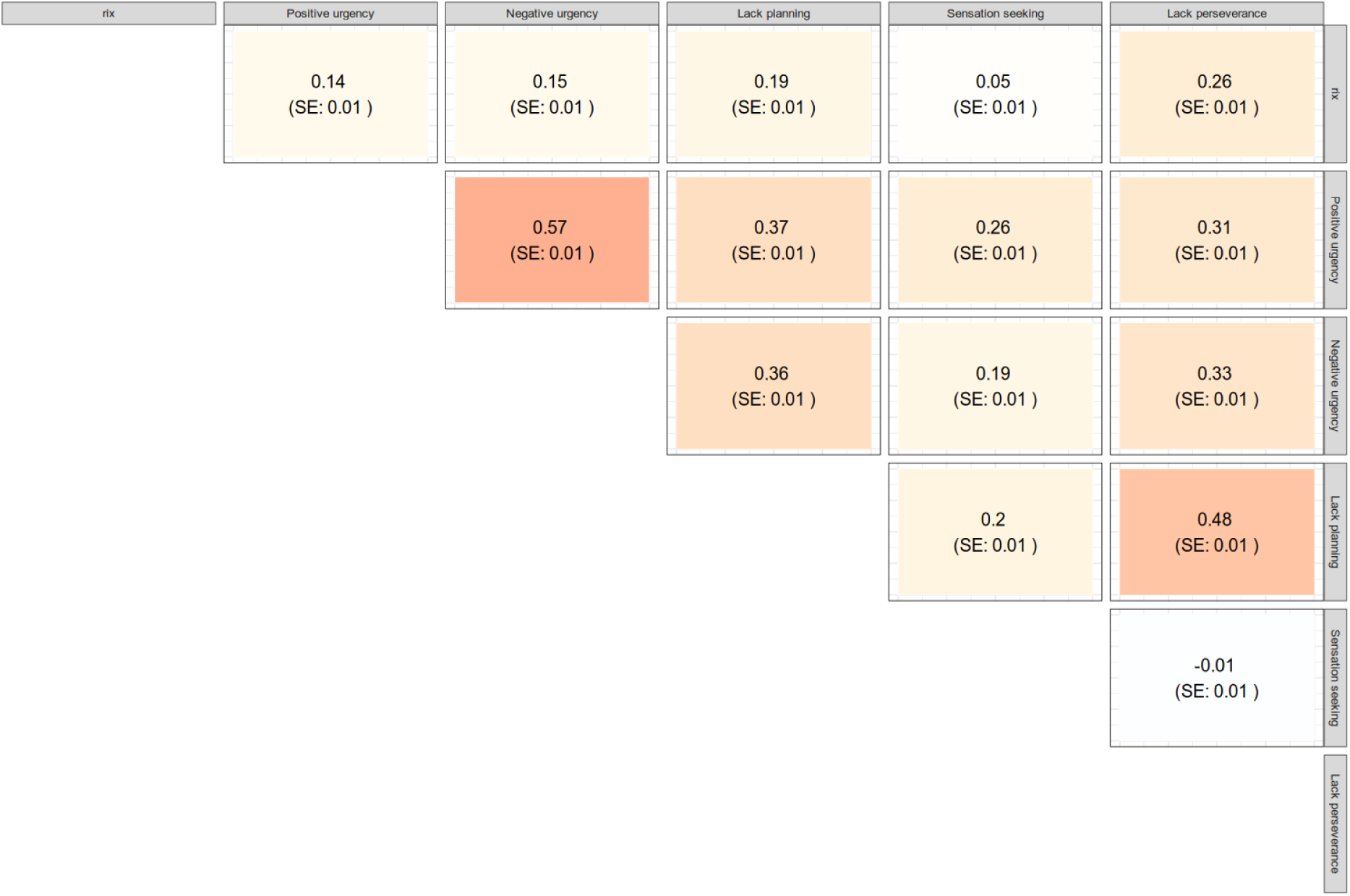
Correlation with impulsivity. Polychoric correlations including the factor score derived from the ADHD symptomatology random intercept (rix, Figure 2) and each impulsivity domain in the UPPS scale treated in the model as ordered ordinals estimated using FIML in the latent likelihood distribution.

### 3.2. Simulations

Simulations were set up for examining the effect of increasing lag on the various parameter estimates. A factorial design exact data simulation (Table 3) was used to obtain parameter estimates as lag increases for CLPM, IV-CLPM, RI-CLPM, and IV-RI-CLPM. Figure 4 through 12 illustrate various aspects of the IV-RI-CLPM and RI-CLPM models to show the internal validity of the new model and changes with regard to power. Data was generated per specification in Figure 1 extended to 20 time-points, once the model reach stationarity a lag was introduced. Figure 4 and Figure 9 are a visual representation of stationarity, or when the cross-sectional correlation between X and Y stop changing. Both models reached stationarity very early in the number of waves, stabilizing at wave ten in the IV-RI-CLPM case (Figure 4) and at wave five in the RI-CLPM case (Figure 9). The two models presented here were only identified with three waves minimum. In order to assess the effect of an increased lag, we designed a simulation procedure where the lag was included between the last and the second-to-last occasion.

### 3.4. Proximal path 1

In the IV-RI-CLPM, as the time between last and second-to-last occasions increased, the proximal 1 path estimate remained unchanged over time. Removing the random intercept did not change this pattern. The particular value where this stability occurs will depend on auto-regressive true values (arp, Figure 5), the variances of the random intercepts (rix parameter, Figure 6), the correlation between random intercepts (rixy, Figure 6), and the innovation variances (u and v, Figure 6); but it is independent of the effect sizes of the instrument (px, in Figure 5 one can see that the gray and yellow lines overlap in the full model and in the model without the random intercept). The presence of the instrument in the model results in the within-wave causal path estimation consistent across time. Importantly, even though the data are generated using an IV-RI-CLPM model, similar IV-RI-CLPMs underestimate the proximal path 1, whereas IV-CLPMs ( without RIs) tend to stay at the true parameter value (true e = [0.1,0.2], Table 3).

**Figure 5.**
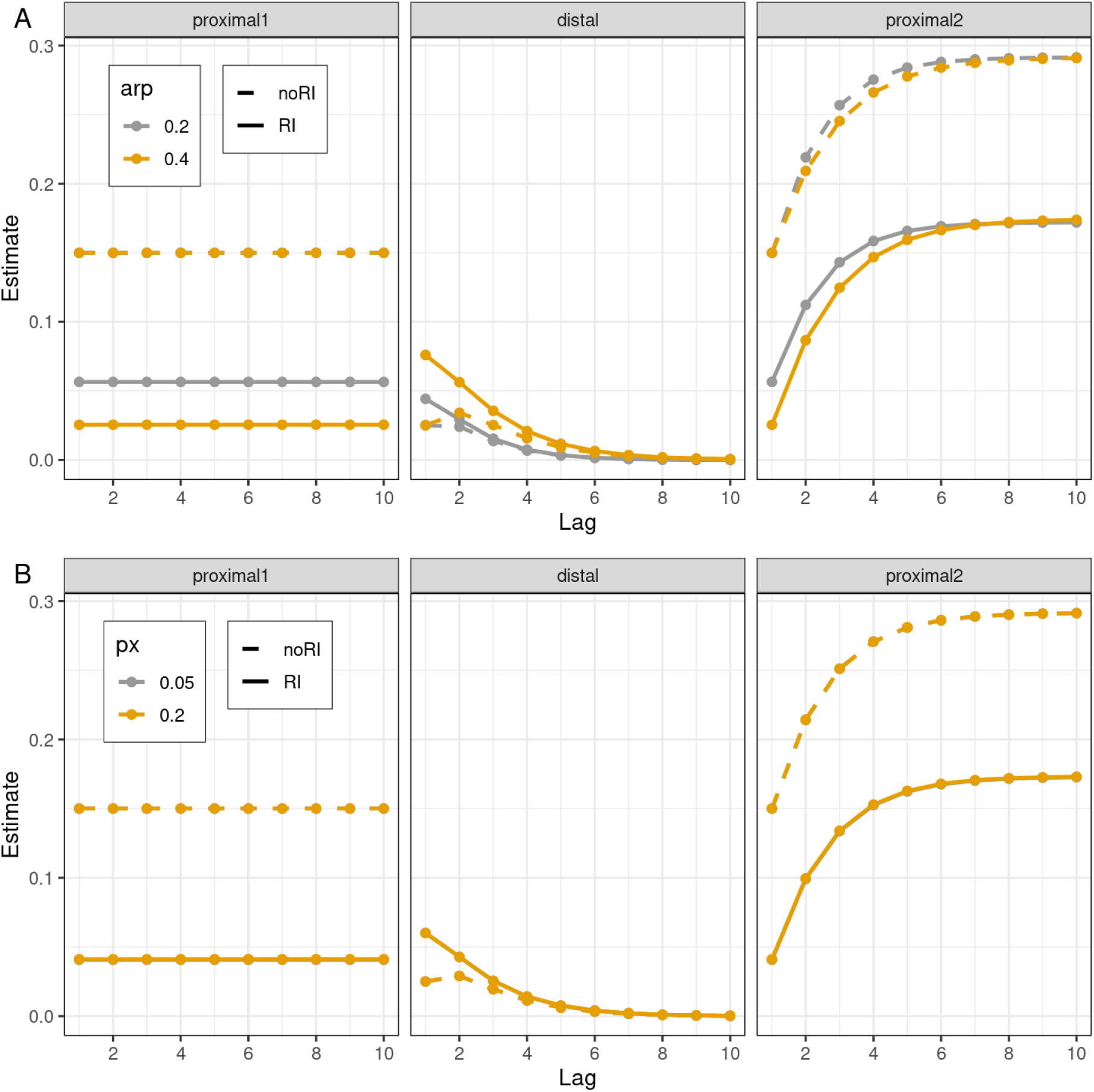
Estimate change in increasing lag - IV-RI-CLPM. After the model reaches stationarity, a lag was simulated. The x-axis represents the length of the lag. The y-axis represents the estimate of the proximal 1, distal or proximal 2 paths in specifications with (RI) and without random intercepts (noRI). NoRI is equivalent to Singh et al. (2023) IV-CLPM. Proximal 1 represents the Xt to Yt effect before the lag is introduced; proximal 2 represents Xt effect on Yt on last occasion. Distal is the cross-lagged path Xt to Yt+lag; a, the path coefficient of the auto-regressive path; px, is the path coefficient of the instrument on x.

**Figure 6:**
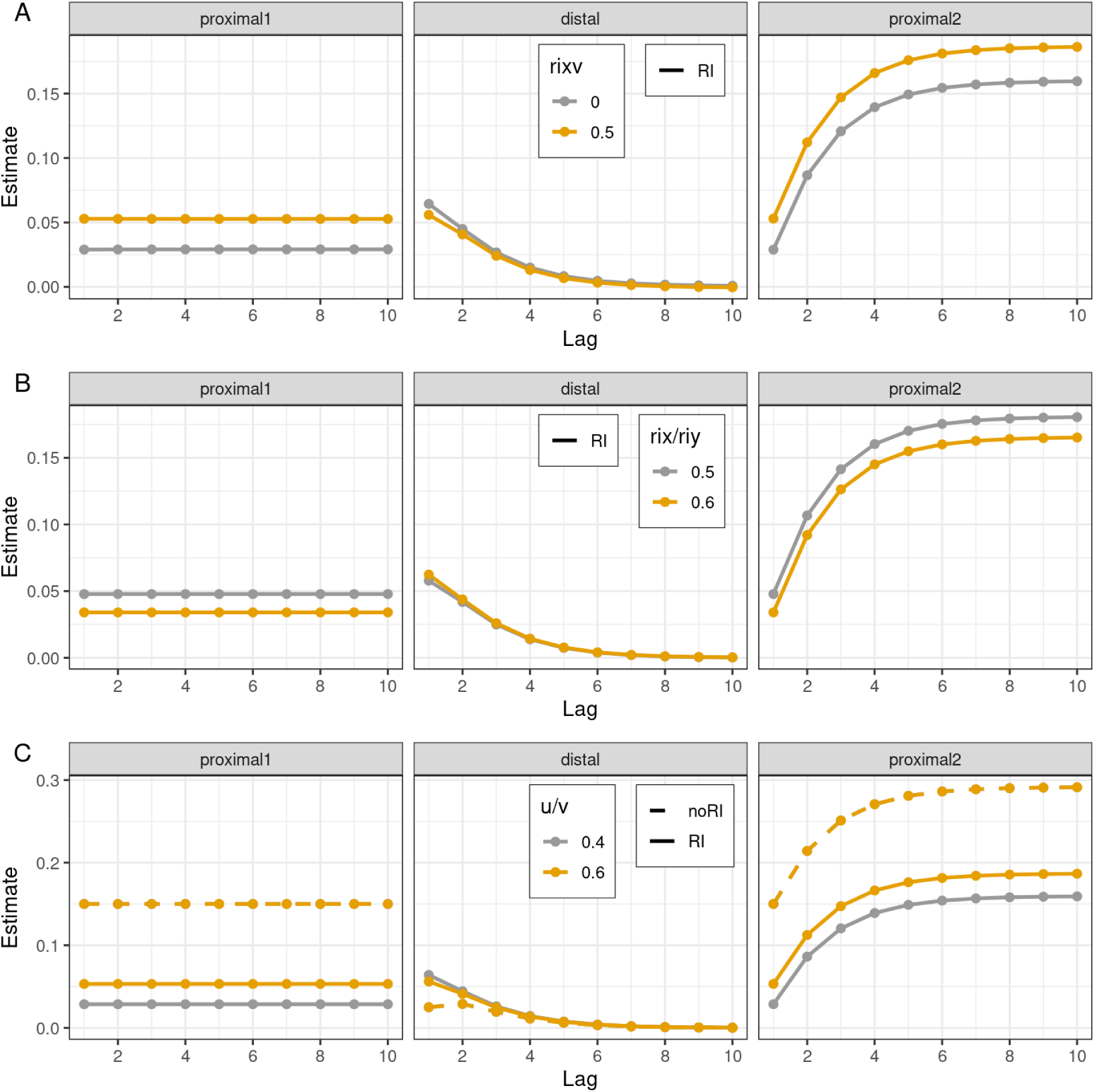
Estimate change over increasing lag - IV-RI-CLPM. After the model reaches stationarity, a lag was simulated. The x-axis represents the length of the lag. The y-axis represents the estimate of the proximal 1, distal or proximal 2 paths in specifications with (RI) and without random intercepts (noRI). NoRI is equivalent to Singh et al. (2023) IV-CLPM. Proximal 1 represents the Xt to Yt effect before the lag is introduced, proximal 2 represents Xt effect on Yt on last occasion. Distal is the cross-lagged path Xt to Yt+lag; r_ri, correlation between the random intercept for X and the RI for Y; RI, the between-person variance (RI variance).

### 3.3. Distal paths (cross-lagged)

Distal path estimate changes over increasing lag. Starting by the IV-RI-CLPM results we can observe that the distal path asymptotes to zero, but the rate in which this happens depends on the effect sizes of the cross-lagged paths (b) and auto-regressive paths (a, Figure 5 and 7). The variances of the random intercepts (rix/riy parameters), the correlation between random intercepts (rixy), the innovation variances (u,v), and the effect size of the instrument for X (px) have a minimal effect on the distal path rate of decay as the lag increases (Figures 3, 4, and 5). A similar pattern is found in the RI-CLPM model (Figure 11), except that the variances of p and q (u/v) affect the rate in which the parameter estimate for the distal path asymptotes to zero.

**Figure 7:**
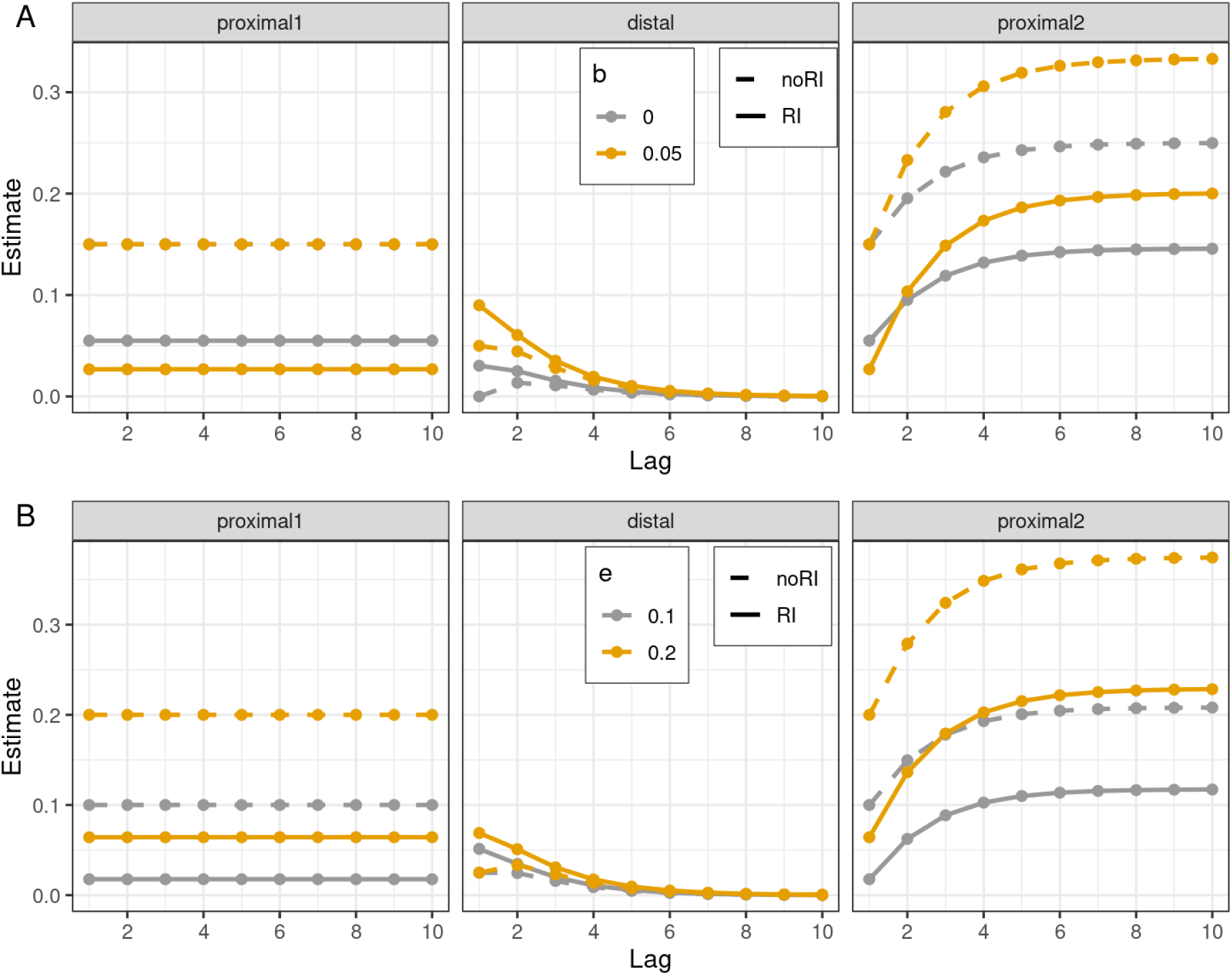
Estimate change over increasing lag - IV-RI-CLPM. After the model reaches stationarity (wave 11), a lag was simulated. The x-axis represents the length of the lag. The y-axis represents the estimate of the proximal 1, distal or proximal 2 paths in specifications with (RI) and without random intercepts (noRI). NoRI is equivalent to Singh et al. (2023) IV-CLPM. Proximal 1 represents the Xt to Yt effect before the lag is introduced, proximal 2 represents Xt effect on Yt on last occasion. Distal is the cross-lagged path Xt to Yt+lag; p2q, the effect size of the cross-lagged path Xt to Yt+1 in the data generation process; immedx, the effect size of the immediate causal path Xt to Yt.

**Figure 10.**
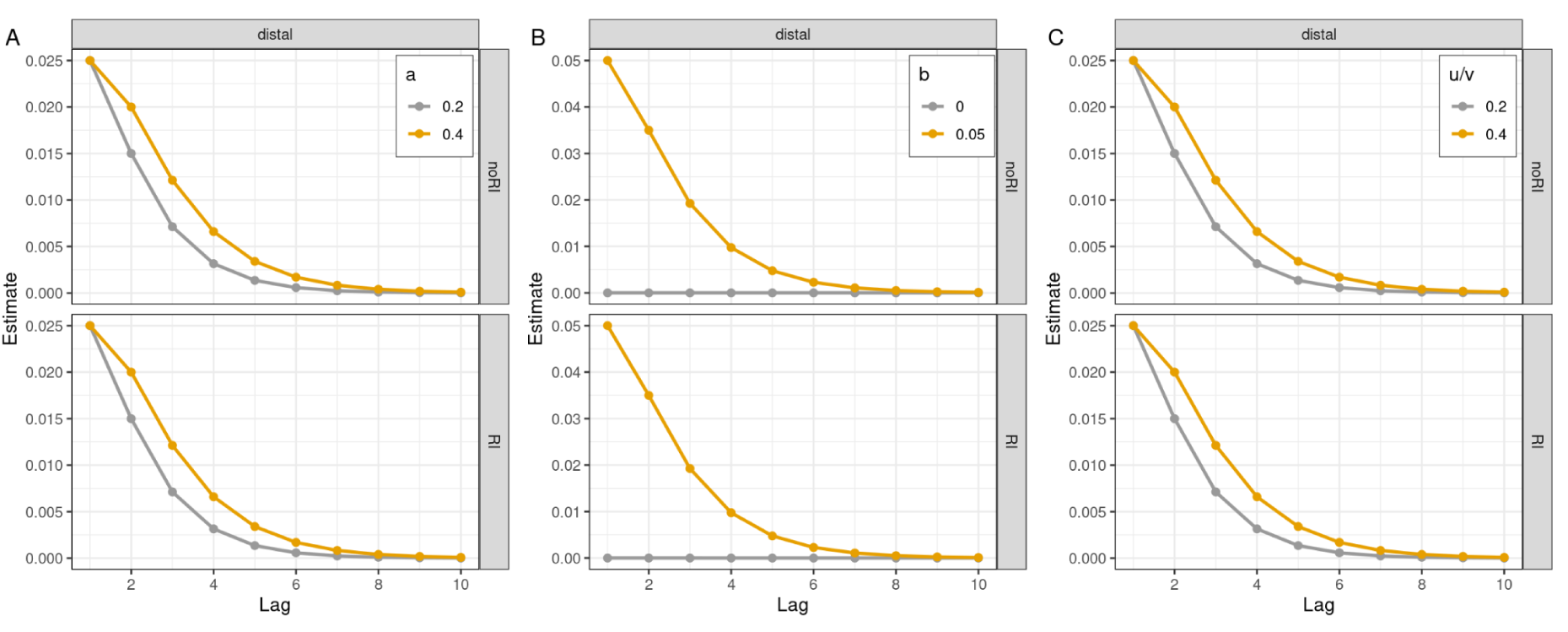
RI-CLPM estimates over increasing lag. After the model reaches stationarity, a lag was simulated. The x-axis represents the length of the lag. The y-axis represents the estimate of the distal path in specifications with (RI) and without random intercepts (noRI). NoRI is equivalent to conventional CLPM. arp, the effect size of the auto-regressive path; u/v, the within-person variance (in RI) or the X and Y variances (in noRI); b, the effect size of the cross-lagged path Xt to Yt+1 in the data generation process.

**Figure 11:**
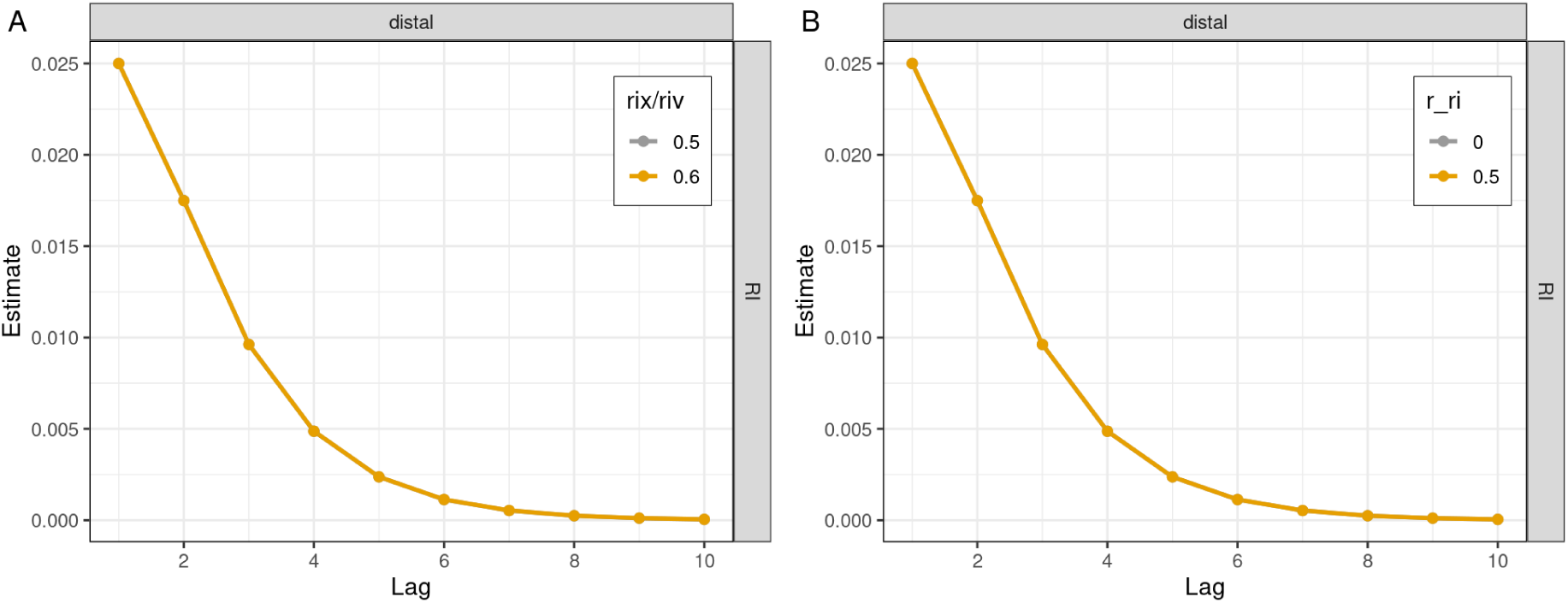
RI-CLPM estimates over increasing lag. After the model reaches stationarity, a lag was simulated. The x-axis represents the length of the lag. The y-axis represents the estimate of the distal path in specifications with (RI) and without random intercepts (noRI). rixy, correlation between the random intercept for X and the RI for Y; xix/riy, the between-person variance (RI variance).

### 3.5. Proximal path 2

As the time between last and second-to-last occasions increase, the proximal 2 path estimate increases. The values of these estimates will vary as a function of auto-regressive true values (a, Figure 5), the variances of the random intercepts (rix/riy, Figure 6), the correlation between random intercepts (rixy, Figure 6), and the innovation variances (u/v, Figure 6); but it is independent of the effect sizes of the instrument (px, Figure 5). Importantly, in the absence of RI (CLPM) models will tend to overestimate the proximal path 2 going above the true values (true e = [0.1,0.2], Table 3) used in the simulation procedure. The proximal paths are also not present in RI-CLPM.

### 3.6. Power

In the case of the IV-RI-CLPM, as the lag increases, the power to detect the distal path decreases (Figure 8). The rate in which the power decreases is a function of the auto-regressive path (a) and the instrument path (px) parameter values. The model specified without a random intercept (IV-CLPM) is overall better powered (Figure 8). In the RI-CLPM case, an identical pattern is revealed. Note that RI-CLPM is less powered to detect the last cross-lagged path given exact same parameter true value used between models (Figure 12).

In summary, the models compared differ in significant ways. All models without RI present better power to detect the distal path (Figure 8 and Figure 12). More importantly, all models’ ability to reject the hypothesis that the distal path is not zero decreases as lag increases. This is consistent with what was found by Singh et al. (2021).

## 4. Discussion

In this study, we employed a novel Instrumental Variable Random Intercept Cross-Lagged Panel Model (IV-RI-CLPM) to investigate the bidirectional relationship between ADHD symptoms and educational performance. Using data from the Adolescent Brain Cognitive Development (ABCD) study, we found significant within-wave bidirectional effects, with ADHD symptoms negatively impacting academic performance and vice versa. The empirical analysis revealed that ADHD symptoms and school grades are relatively stable constructs over time, with significant auto-regressive paths. Interestingly, the within-wave causal paths were consistently negative, underscoring the immediate detrimental effects of ADHD symptoms on academic performance and vice versa. One cross-lagged path from ADHD symptoms in flw3 to education performance in flw4 was significant (b2 = −0.1). Simulation studies validated our model, demonstrating its robustness in capturing within-wave causal relations, and revealed that CLPM models with or without RI are biased when lag increases. This is consistent with the findings from Singh et al. (2023) using classic CLPM and IV-CLPM.

Our findings align with previous research indicating a bidirectional relationship between ADHD and educational outcomes (Castro-de-Araujo et al., 2024; Demange et al., 2024). Castro-de-Araujo et al., (2024) found an effect size for the ADHD to EP causal path (g1) −0.268 [95%CI −0.412, −0.124] (estimated −0.189 [ 95%CI −0.273, −0.104] in the IV-RI-CLPM) and EP to ADHD causal path (g2) −0.731 [95%CI −1.11, −0.35] ( −0.183 [95%CI −0.309, −0.06] path in the IV-RI-CLPM ). However, MR-DoC2 estimates family-level causal effect, whereas IV-RI-CLPM estimates a within-individual effect of the state-like variation. IV-RI-CLPM provides a more nuanced view of the temporal dynamics involved between these phenotypes. Contrary to our initial hypothesis, no consistent cross-lagged causal effects were found. This might be due, as it was shown through simulations, that CLPM models in general struggle to return unbiased estimates when lag increases.

The significant within-wave effects suggest that interventions targeting ADHD symptoms are likely to have immediate benefits for academic performance and vice versa. This highlights the importance of integrated approaches that address both academic and behavioral aspects in individuals with ADHD. The reduction in the instrument effects over time in the empirical analysis suggests that genetic influences may diminish as environmental factors become more prominent, particularly for ADHD scores.

The simulation studies provided valuable insights into the model’s performance under numerous conditions. IV-RI-CLPM maintain stable proximal path 1 estimates despite increasing lag. The proximal path 2 summarizes accrued causal effects as the lag increases in IV-RI-CLPM. On the other hand, the distal path shows a progressive downward bias. As time between measurement increases, the estimated cross-lagged effect diminishes as the model’s power to detect causal effects, consistent with theoretical expectations (Singh et al., 2023). This explains a lack of consistent cross-lagged effect in the substantive application using ADHD and EP.

These results should be considered in light of some limitations. First, study wave four was incomplete at the time of analysis, which may have introduced selection bias and biases from non-random missingness, which is a required assumption by the estimation method. Second, we only analyzed data from individuals of European ancestry, hindering this analysis generalizability. Third, it is possible that the autoregressive and cross-lagged paths do not explain the unobserved influences or time-specific variance at each variable, leading to omitted variable bias. However, specific variances that are not transmitted can be included in this model provided data for five waves are available. Fourth, IV-RI-CLPM does not include horizontal pleiotropy, and its presence certainly bias the causal paths. Finally, it should be noted that, to be consistent with IV-CLPM, the instruments only predict the state-level fluctuations in the model - not the RI (the stable trait).

These findings have important implications for clinical practice and educational policy. The effect of education performance on ADHD might be resulting from parents changing the home environment to adapt to their children education deficiencies, providing extra support, which in turn might have an immediate effect on ADHD symptomatology. Future research should attempt to model horizontal pleiotropy and a direct effect of the instrumental variables on random intercepts, thus controlling for genetic effects on the trait-like variation.

## Declarations

### Ethics approval

Not applicable

### Consent for publication

Not applicable

### Availability of data and material

Data used in the preparation of this article were obtained from the Adolescent Brain Cognitive Development (ABCD) Study (https://abcdstudy.org), held in the NIMH Data Archive (NDA). This is a multisite, longitudinal study designed to recruit more than 10,000 children age 9-10 and follow them over 10 years into early adulthood. A listing of participating sites and a complete listing of the study investigators can be found at https://abcdstudy.org/consortium_members/.

### Code availability

Scripts are available in a repository for replication (https://github.com/lf-araujo/2024-RI-CLPM). The scripts used for genetic ancestry assignment and genetic principal component analysis are available upon request. An optimized function for the IV-RI-CLPM is available in the R umx package (Bates et al., 2019)

### Author contributions

ABCD consortium investigators designed and implemented the study and/or provided data but, apart from Dr. Neale, did not participate in the analysis or writing of this report. This manuscript reflects the views of the authors and may not reflect the opinions or views of the NIH or ABCD consortium investigators. LA performed all analyses. SK, supported in the analyses. DZ calculated the PS. All authors contributed in writing the manuscript.

## References

Achenbach, T.M., 2013. DSM-Oriented Guide for the Achenbach System of Empirically Based Assessment (ASEBA®).

Achenbach, T.M., 2001. Manual for ASEBA school-age forms & profiles. Univ. Vt. Res. Cent. Child. Youth Fam.

APA, 2013. Diagnostic and statistical manual of mental disorders (DSM-5), 5th ed. Washington, DC.

Bates, T.C., Maes, H., Neale, M.C., 2019. umx: Twin and Path-Based Structural Equation Modeling in R. Twin Res. Hum. Genet. 22, 27–41. 10.1017/thg.2019.2

Boomsma, D.I., Saviouk, V., Hottenga, J.-J., Distel, M.A., de Moor, M.H.M., Vink, J.M., Geels, L.M., van Beek, J.H.D.A., Bartels, M., de Geus, E.J.C., Willemsen, G., 2010. Genetic Epidemiology of Attention Deficit Hyperactivity Disorder (ADHD Index) in Adults. PLoS ONE 5, e10621. 10.1371/journal.pone.0010621

Branigan, A.R., McCallum, K.J., Freese, J., 2013. Variation in the Heritability of Educational Attainment: An International Meta-Analysis. Soc. Forces 92, 109–140. 10.1093/sf/sot076

Castro-de-Araujo, L., Singh, M., Zhou, Y., Vinh, P., Maes, H.H.M., Verhulst, B., Dolan, C., Neale, M.C., 2023. Power, measurement error, and pleiotropy robustness in twin-design extensions to Mendelian Randomization. 10.21203/rs.3.rs-3411642/v1

Castro-de-Araujo, L.F., Zhou, D. (Yi), Su, M.-H., Kramer, S., Loughnan, R., Neale, M.C., 2024. Quasi-experimental analyses of the effect of ADHD on education performance in youths across sexes and ancestry. 10.21203/rs.3.rs-4638880/v1

Chang, L.-Y., Wang, M.-Y., Tsai, P.-S., 2016. Diagnostic Accuracy of Rating Scales for Attention-Deficit/Hyperactivity Disorder: A Meta-analysis. Pediatrics 137, e20152749. 10.1542/peds.2015-2749

Chen, W., Zhou, K., Sham, P., Franke, B., Kuntsi, J., Campbell, D., Fleischman, K., Knight, J., Andreou, P., Arnold, R., Altink, M., Boer, F., Boholst, M.J., Buschgens, C., Butler, L., Christiansen, H., Fliers, E., Howe-Forbes, R., Gabriëls, I., Heise, A., Korn-Lubetzki, I., Marco, R., Medad, S., Minderaa, R., Müller, U.C., Mulligan, A., Psychogiou, L., Rommelse, N., Sethna, V., Uebel, H., McGuffin, P., Plomin, R., Banaschewski, T., Buitelaar, J., Ebstein, R., Eisenberg, J., Gill, M., Manor, I., Miranda, A., Mulas, F., Oades, R.D., Roeyers, H., Rothenberger, A., Sergeant, J., Sonuga-Barke, E., Steinhausen, H.-C., Taylor, E., Thompson, M., Faraone, S.V., Asherson, P., 2008. DSM-IV combined type ADHD shows familial association with sibling trait scores: a sampling strategy for QTL linkage. Am. J. Med. Genet. Part B Neuropsychiatr. Genet. Off. Publ. Int. Soc. Psychiatr. Genet. 147B, 1450–1460. 10.1002/ajmg.b.30672

Choi, W.-S., Woo, Y.S., Wang, S.-M., Lim, H.K., Bahk, W.-M., 2022. The prevalence of psychiatric comorbidities in adult ADHD compared with non-ADHD populations: A systematic literature review. PLOS ONE 17, e0277175. 10.1371/journal.pone.0277175

Conomos, M.P., Miller, M.B., Thornton, T.A., 2015. Robust inference of population structure for ancestry prediction and correction of stratification in the presence of relatedness. Genet. Epidemiol. 39, 276–293. 10.1002/gepi.21896

Dardani, C., Riglin, L., Leppert, B., Sanderson, E., Rai, D., Howe, L.D., Davey Smith, G., Tilling, K., Thapar, A., Davies, N.M., Anderson, E., Stergiakouli, E., 2021. Is genetic liability to ADHD and ASD causally linked to educational attainment? Int. J. Epidemiol. 50, 2011–2023. 10.1093/ije/dyab107

Demange, P.A., Boomsma, D.I., van Bergen, E., Nivard, M.G., 2024. Educational attainment and psychiatric diagnoses: a national registry data and two-sample Mendelian randomization study. Nat. Ment. Health 2, 668–679. 10.1038/s44220-024-00245-x

Demange, P.A., Boomsma, D.I., van Bergen, E., Nivard, M.G., 2023. Evaluating the causal relationship between educational attainment and mental health. medRxiv 2023.01.26.23285029. 10.1101/2023.01.26.23285029

Demontis, D., Walters, R.K., Martin, J., Mattheisen, M., Als, T.D., Agerbo, E., Baldursson, G., Belliveau, R., Bybjerg-Grauholm, J., Bækvad-Hansen, M., Cerrato, F., Chambert, K., Churchhouse, C., Dumont, A., Eriksson, N., Gandal, M., Goldstein, J.I., Grasby, K.L., Grove, J., Gudmundsson, O.O., Hansen, C.S., Hauberg, M.E., Hollegaard, M.V., Howrigan, D.P., Huang, H., Maller, J.B., Martin, A.R., Martin, N.G., Moran, J., Pallesen, J., Palmer, D.S., Pedersen, C.B., Pedersen, M.G., Poterba, T., Poulsen, J.B., Ripke, S., Robinson, E.B., Satterstrom, F.K., Stefansson, H., Stevens, C., Turley, P., Walters, G.B., Won, H., Wright, M.J., Consortium (PGC), A.W.G. of the P.G., Consortium, E.L.& G.E. (EAGLE), Team, 23 and Me Research, Andreassen, O.A., Asherson, P., Burton, C.L., Boomsma, D.I., Cormand, B., Dalsgaard, S., Franke, B., Gelernter, J., Geschwind, D., Hakonarson, H., Haavik, J., Kranzler, H.R., Kuntsi, J., Langley, K., Lesch, K.-P., Middeldorp, C., Reif, A., Rohde, L.A., Roussos, P., Schachar, R., Sklar, P., Sonuga-Barke, E.J.S., Sullivan, P.F., Thapar, A., Tung, J.Y., Waldman, I.D., Medland, S.E., Stefansson, K., Nordentoft, M., Hougaard, D.M., Werge, T., Mors, O., Mortensen, P.B., Daly, M.J., Faraone, S.V., Børglum, A.D., Neale, B.M., 2019. Discovery of the first genome-wide significant risk loci for attention-deficit/hyperactivity disorder. Nat. Genet. 51, 63. 10.1038/s41588-018-0269-7

Dick, A.S., Lopez, D.A., Watts, A.L., Heeringa, S., Reuter, C., Bartsch, H., Fan, C.C., Kennedy, D.N., Palmer, C., Marshall, A., Haist, F., Hawes, S., Nichols, T.E., Barch, D.M., Jernigan, T.L., Garavan, H., Grant, S., Pariyadath, V., Hoffman, E., Neale, M., Stuart, E.A., Paulus, M.P., Sher, K.J., Thompson, W.K., 2021. Meaningful associations in the adolescent brain cognitive development study. NeuroImage 239, 118262. 10.1016/j.neuroimage.2021.118262

Faraone, S.V., Larsson, H., 2019. Genetics of attention deficit hyperactivity disorder. Mol. Psychiatry 24, 562–575. 10.1038/s41380-018-0070-0

Fayyad, J., Sampson, N.A., Hwang, I., Adamowski, T., Aguilar-Gaxiola, S., Al-Hamzawi, A., Andrade, L.H.S.G., Borges, G., de Girolamo, G., Florescu, S., Gureje, O., Haro, J.M., Hu, C., Karam, E.G., Lee, S., Navarro-Mateu, F., O’Neill, S., Pennell, B.-E., Piazza, M., Posada-Villa, J., Have, M. ten, Torres, Y., Xavier, M., Zaslavsky, A.M., Kessler, R.C., 2017. The descriptive epidemiology of DSM-IV Adult ADHD in the World Health Organization World Mental Health Surveys. Atten. Deficit Hyperact. Disord. 9, 47–65. 10.1007/s12402-016-0208-3

Ge, T., Chen, C.-Y., Ni, Y., Feng, Y.-C.A., Smoller, J.W., 2019. Polygenic prediction via Bayesian regression and continuous shrinkage priors. Nat. Commun. 10, 1776. 10.1038/s41467-019-09718-5

Haist, F., Jernigan, T.L., 2023. Adolescent Brain Cognitive Development Study (ABCD) - Annual Release 5.0. 10.15154/8873-ZJ65

Hamaker, E.L., Kuiper, R.M., Grasman, R.P.P.P., 2015. A critique of the cross-lagged panel model. Psychol. Methods 20, 102–116. 10.1037/a0038889

Hinshaw, S.P., Nguyen, P.T., O’Grady, S.M., Rosenthal, E.A., 2022. Annual Research Review: Attention-deficit/hyperactivity disorder in girls and women: underrepresentation, longitudinal processes, and key directions. J. Child Psychol. Psychiatry 63, 484–496. 10.1111/jcpp.13480

Hunter, M.D., Garrison, S.M., Burt, S.A., Rodgers, J.L., 2021. The Analytic Identification of Variance Component Models Common to Behavior Genetics. Behav. Genet. 51, 425–437. 10.1007/s10519-021-10055-x

Jiang, Z., Wang, D., Xu, H., Zhang, A., Zhao, Q., Yan, J., Wang, X., Zhang, W., Li, Yanlin, Yang, K., Hu, S., Cui, Y., Li, Ying, 2023. Diagnostic efficiency and psychometric properties of CBCL DSM-oriented scales in a large sample of Chinese school-attending students aged 5-16. Asian J. Psychiatry 88, 103724. 10.1016/j.ajp.2023.103724

Kenny, D.A., 1975. Cross-lagged panel correlation: A test for spuriousness. Psychol. Bull. 82, 887–903. 10.1037/0033-2909.82.6.887

Lahey, B.B., Tong, L., Pierce, B., Hedeker, D., Berman, M.G., Cardenas-Iniguez, C., Moore, T.M., Applegate, B., Tiemeier, H., Kaczkurkin, A.N., 2022. Associations of polygenic risk for attention-deficit/hyperactivity disorder with general and specific dimensions of childhood psychological problems and facets of impulsivity. J. Psychiatr. Res. 152, 187–193. 10.1016/j.jpsychires.2022.06.019

Larsson, H., Chang, Z., D’Onofrio, B.M., Lichtenstein, P., 2014. The heritability of clinically diagnosed attention deficit hyperactivity disorder across the lifespan. Psychol. Med. 44, 2223–2229. 10.1017/S0033291713002493

Lee, J.J., Wedow, R., Okbay, A., Kong, E., Maghzian, O., Zacher, M., Nguyen-Viet, T.A., Bowers, P., Sidorenko, J., Karlsson Linnér, R., Fontana, M.A., Kundu, T., Lee, C., Li, H., Li, R., Royer, R., Timshel, P.N., Walters, R.K., Willoughby, E.A., Yengo, L., Alver, M., Bao, Y., Clark, D.W., Day, F.R., Furlotte, N.A., Joshi, P.K., Kemper, K.E., Kleinman, A., Langenberg, C., Mägi, R., Trampush, J.W., Verma, S.S., Wu, Y., Lam, M., Zhao, J.H., Zheng, Z., Boardman, J.D., Campbell, H., Freese, J., Harris, K.M., Hayward, C., Herd, P., Kumari, M., Lencz, T., Luan, J., Malhotra, A.K., Metspalu, A., Milani, L., Ong, K.K., Perry, J.R.B., Porteous, D.J., Ritchie, M.D., Smart, M.C., Smith, B.H., Tung, J.Y., Wareham, N.J., Wilson, J.F., Beauchamp, J.P., Conley, D.C., Esko, T., Lehrer, S.F., Magnusson, P.K.E., Oskarsson, S., Pers, T.H., Robinson, M.R., Thom, K., Watson, C., Chabris, C.F., Meyer, M.N., Laibson, D.I., Yang, J., Johannesson, M., Koellinger, P.D., Turley, P., Visscher, P.M., Benjamin, D.J., Cesarini, D., 2018. Gene discovery and polygenic prediction from a genome-wide association study of educational attainment in 1.1 million individuals. Nat. Genet. 50, 1112–1121. 10.1038/s41588-018-0147-3

Maes, H.H.M., Lapato, D.M., Schmitt, J.E., Luciana, M., Banich, M.T., Bjork, J.M., Hewitt, J.K., Madden, P.A., Heath, A.C., Barch, D.M., Thompson, W.K., Iacono, W.G., Neale, M.C., 2022. Genetic and Environmental Variation in Continuous Phenotypes in the ABCD Study®. Behav. Genet. 10.1007/s10519-022-10123-w

Michaëlsson, M., Yuan, S., Melhus, H., Baron, J.A., Byberg, L., Larsson, S.C., Michaëlsson, K., 2022. The impact and causal directions for the associations between diagnosis of ADHD, socioeconomic status, and intelligence by use of a bi-directional two-sample Mendelian randomization design. BMC Med. 20, 106. 10.1186/s12916-022-02314-3

Murray, C.J.L., 2013. The State of US Health, 1990-2010: Burden of Diseases, Injuries, and Risk Factors. JAMA 310, 591. 10.1001/jama.2013.13805

Neale, M.C., Hunter, M.D., Pritikin, J.N., Zahery, M., Brick, T.R., Kirkpatrick, R.M., Estabrook, R., Bates, T.C., Maes, H.H., Boker, S.M., 2016. OpenMx 2.0: Extended Structural Equation and Statistical Modeling. Psychometrika 81, 535–549. 10.1007/s11336-014-9435-8

Owens, M.M., Potter, A., Hyatt, C.S., Albaugh, M., Thompson, W.K., Jernigan, T., Yuan, D., Hahn, S., Allgaier, N., Garavan, H., 2021. Recalibrating expectations about effect size: A multi-method survey of effect sizes in the ABCD study. PLoS ONE 16, e0257535. 10.1371/journal.pone.0257535

Purcell, S., Neale, B., Todd-Brown, K., Thomas, L., Ferreira, M.A.R., Bender, D., Maller, J., Sklar, P., de Bakker, P.I.W., Daly, M.J., Sham, P.C., 2007. PLINK: A Tool Set for Whole-Genome Association and Population-Based Linkage Analyses. Am. J. Hum. Genet. 81, 559–575. 10.1086/519795

Rimfeld, K., Krapohl, E., Trzaskowski, M., Coleman, J.R.I., Selzam, S., Dale, P.S., Esko, T., Metspalu, A., Plomin, R., 2018. Genetic influence on social outcomes during and after the Soviet era in Estonia. Nat. Hum. Behav. 2, 269–275. 10.1038/s41562-018-0332-5

Sanchez-Roige, S., Jennings, M.V., Thorpe, H.H.A., Mallari, J.E., van der Werf, L.C., Bianchi, S.B., Huang, Y., Lee, C., Mallard, T.T., Barnes, S.A., Wu, J.Y., Barkley-Levenson, A.M., Boussaty, E.C., Snethlage, C.E., Schafer, D., Babic, Z., Winters, B.D., Watters, K.E., Biederer, T., Mackillop, J., Stephens, D.N., Elson, S.L., Fontanillas, P., Khokhar, J.Y., Young, J.W., Palmer, A.A., 2023. CADM2 is implicated in impulsive personality and numerous other traits by genome- and phenome-wide association studies in humans and mice. Transl. Psychiatry 13, 1–11. 10.1038/s41398-023-02453-y

Selig, J.P., Little, T.D., 2012. Autoregressive and cross-lagged panel analysis for longitudinal data, in: Handbook of Developmental Research Methods. The Guilford Press, New York, NY, US, pp. 265–278.

Singh, M., Dolan, C.V., Neale, M.C., 2023. Integrating Cross-Lagged Panel Models with Instrumental Variables to Extend the Temporal Generalizability of Causal Inference. Multivar. Behav. Res. 1–2. 10.1080/00273171.2022.2160954

Sousa, A. de F., Isabel, C., Castanho, J., Polanczyk, G., Rohde, L., 2020. Attention-defficit hyperactivity disorders, in: Rey, J. (Ed.), E-Textbook of Child and Adolescent Mental Health. International Association for Child and Adolescent Psychiatry and Allied Professions, Geneva.

Sprich, S., Biederman, J., Crawford, M.H., Mundy, E., Faraone, S.V., 2000. Adoptive and biological families of children and adolescents with ADHD. J. Am. Acad. Child Adolesc. Psychiatry 39, 1432–1437. 10.1097/00004583-200011000-00018

Taylor, M.J., Lichtenstein, P., Larsson, H., Anckarsäter, H., Greven, C.U., Ronald, A., 2016. Is There a Female Protective Effect Against Attention-Deficit/Hyperactivity Disorder? Evidence From Two Representative Twin Samples. J. Am. Acad. Child Adolesc. Psychiatry 55, 504–512.e2. 10.1016/j.jaac.2016.04.004

Thomas, R., Sanders, S., Doust, J., Beller, E., Glasziou, P., 2015. Prevalence of attention-deficit/hyperactivity disorder: a systematic review and meta-analysis. Pediatrics 135, e994–1001. 10.1542/peds.2014-3482

van der Sluis, S., Dolan, C.V., Neale, M.C., Posthuma, D., 2008. Power Calculations Using Exact Data Simulation: A Useful Tool for Genetic Study Designs. Behav. Genet. 38, 202–211. 10.1007/s10519-007-9184-x

Venables, W.N., Ripley, B.D., Venables, W.N., 2002. Modern applied statistics with S, 4th ed. ed, Statistics and computing. Springer, New York.

Willcutt, E.G., 2012. The prevalence of DSM-IV attention-deficit/hyperactivity disorder: a meta-analytic review. Neurother. J. Am. Soc. Exp. Neurother. 9, 490–499. 10.1007/s13311-012-0135-8

